# Topological Deep Learning Identifies Polygenic Variant Clusters Across Familial Multimorbid Disorders

**DOI:** 10.64898/2026.06.03.26354242

**Authors:** Kelly Larissa Vomo-Donfack, Guilhem Bousquet, Géraldine Falgarone, Grégory Ginot, Ian Morilla

## Abstract

Whole-genome sequencing comprehensively captures coding, non-coding and structural variation in families with suspected inherited disorders, yet its clinical utility remains constrained by an interpretation bottleneck: selecting a handful of relevant variants from millions of candidates. Current rule-based pipelines, anchored in ACMG/AMP criteria, excel at identifying highly penetrant Mendelian alleles but frequently miss variants of low-to-moderate penetrance, non-coding alterations and germline–somatic interactions. Here we introduce PolyCLIP-T, a topology-guided multimodal framework that transforms variant selection from a classification problem into a geometric discovery task. By contrastively aligning DNA-sequence embeddings with functional annotations, PolyCLIP-T constructs a unified latent space in which the displacement between reference and alternate embeddings quantifies the molecular perturbation induced by each variant. Persistent homology then identifies stable topological components— coherent variant groups shared among affected relatives—that transcend single-variant scoring logic. Applied to six families with multi-morbid cancer, autoimmune and cardiovascular disease, PolyCLIP-T recovered non-coding and structural candidates overlooked by conventional pipelines and revealed pleiotropic networks spanning disease categories. This approach provides an interpretable, scalable solution for genome-first investigations of disorders driven by polygenic architectures that evade single-variant analysis. The framework was developed and benchmarked on deeply characterised familial cohorts selected for transgenerational multimorbidity; validation in larger, independent populations will be essential to establish its generalisability. An interactive web tool is freely available at https://www.polyclip-t.uma.es/.

## Introduction

Whole-genome sequencing (WGS) has transformed clinical genetics by offering a single assay that surveys the vast majority of the *∼*3.2 billion base pairs of the human genome[1, 2]. This near-complete coverage enables simultaneous detection of single-nucleotide variants (SNVs), indels, copy-number variants (CNVs), balanced structural rearrangements and complex genomic alterations— collectively providing unprecedented access to the regulatory architecture and structural complexity that governs genome integrity[3]. Consequently, WGS consistently improves diagnostic yield over exome-based strategies by 10–35%, primarily through identification of pathogenic non-coding variants, complex structural variants and regions with poor exome capture efficiency[2, 4].

Despite this transformative potential, widespread clinical implementation of WGS is constrained not by technological limitations but by a profound interpretation bottleneck. Each individual genome harbours 4–5 million variants relative to the reference sequence, of which only a vanishingly small fraction can be attributed to a patient’s phenotype. This prioritisation problem scales exponentially in familial studies, where data from multiple affected and unaffected relatives must be integrated to discern meaningful segregation patterns. Current bioinformatic pipelines, built largely upon the American College of Medical Genetics and Genomics and the Association for Molecular Pathology (ACMG/AMP) guidelines[5], filter variants based on population frequency, predicted functional impact, and mode of inheritance. Yet these pipelines overwhelmingly produce lengthy lists of candidates—many classified as variants of uncertain significance (VUS)—leaving clinicians with an intractable interpretation burden[1, 6]. A major contributor to this difficulty is that a substantial proportion of these VUS likely possess functional consequences that remain unrecognised, as our understanding of the non-coding genome, regulatory elements, and variant effect mechanisms is still far from complete.

The ACMG/AMP framework provides essential rigour and consistency for single-variant interpretation in highly penetrant Mendelian disorders. However, its rule-based, categorical logic is fundamentally ill-suited for the genome-wide data deluge from WGS and, more critically, for identifying variants that contribute to disease through quantitative, combinatorial mechanisms. This includes variants with low or incomplete penetrance, where an allele increases disease risk but does not guarantee it, and oligogenic or polygenic models, where disease manifests from the cumulative burden or interaction of multiple alleles across the genome[7]. These limitations are magnified when considering non-coding variation: despite WGS providing comprehensive data on intronic and intergenic regions, most clinical analyses remain focused on coding exons due to the profound difficulty of assigning pathogenicity to regulatory variants[8]. The functional impact of a non-coding variant depends on a complex, context-dependent interplay of factors—its position relative to cis-regulatory elements, its effect on transcription factor binding motifs, its potential to disrupt splice sites or alter RNA stability, and its role within topologically associating domains (TADs) that orchestrate long-range gene regulation[9]. Deep-learning models have achieved remarkable success in predicting splice-altering variants[10] or chromatin effects[11], yet these computational predictions constitute only moderate-strength evidence (PP3/BP4 criteria) within the ACMG/AMP framework and are challenging to integrate with orthogonal data types.

These collective limitations become critically important for a broad class of familial disorders where familial aggregation is clear yet a single, highly penetrant Mendelian mutation remains elusive—a scenario frequently encountered in familial cancer syndromes in which conventional single-gene testing fails to identify a causative variant[12, 13]. In many cancers, autoimmune conditions and cardiovascular diseases, the genetic architecture can involve variants of low to moderate penetrance acting in combination. Disease may arise from a combination of several risk alleles within the same biological pathway (an oligogenic model); a germline predisposition variant requiring a somatic second hit (as in hereditary cancer syndromes); or the cumulative burden of common variants aggregated into a polygenic risk score[14]. In such cases, a substantial proportion of the contributing variants may not meet classical criteria for pathogenicity: they may be insufficiently rare in population databases to satisfy the ‘very low frequency’ criterion (PM2), their individual effect sizes in functional assays may be modest, and their segregation with disease may be partial rather than complete. A variant-by-variant scoring system that evaluates each allele in isolation against binary pathogenic–benign thresholds is therefore not well suited to detecting the collective biological signal that could emerge from a constellation of such variants. What is needed is a shift in focus from classification to selection—a method that identifies coherent sets of variants that, together, point to a plausible disease mechanism shared among affected family members.

To address this unmet need, we introduce PolyCLIP-T, a topology-guided multimodal framework specifically engineered for the selection of variant sets in familial WGS analysis (Fig. 1). PolyCLIPT is designed to learn multimodal representations of genomic variants by aligning sequence, biological, and text-derived information within a shared embedding space, using ontology-based semantic similarity as a soft-label supervision strategy. Rather than relying on hard pathogenicity labels—which are scarce, biased towards coding regions, and subject to the circularities discussed above—the model learns to capture functional similarity and disease-related relationships between variants through contrastive multimodal learning. In particular, PolyCLIP-T leverages genomic sequence context together with biological annotations and ontology-derived semantic relationships to model the complex, polygenic mechanisms involved in chronic diseases where multiple variants of low-to-moderate penetrance act in combination across cancer, autoimmune, and cardiovascular phenotypes. This design reflects the central hypothesis of the study: that variants contributing to familial multimorbidity do not act in isolation but participate in coherent biological programmes that can be recovered through topological analysis of a jointly learned embedding space. Our objective is not to replace the ACMG/AMP framework for definitive variant classification but to operate upstream of it, providing a powerful filtering and prioritisation layer that identifies the small subset of variants most worthy of detailed, manual curation. PolyCLIP-T integrates heterogeneous data streams—raw DNA sequence, evolutionary conservation, functional genomic annotations and population genetics—into a unified mathematical representation. A contrastive learning model aligns reference and alternate DNA-sequence embeddings with high-dimensional representations of their functional consequences, creating a semantically rich feature space for each variant[15, 16]. We then apply topological data analysis (TDA)[17], specifically persistent homology, to this integrated feature space to identify persistent topological components—clusters or connected structures in the data space that remain stable across multiple scales. These persistent components represent coherent groups of variants that are functionally similar and shared across related affected individuals. Variants that together form a strong, persistent topological signal are prioritised for downstream evaluation.

**Figure 1:**
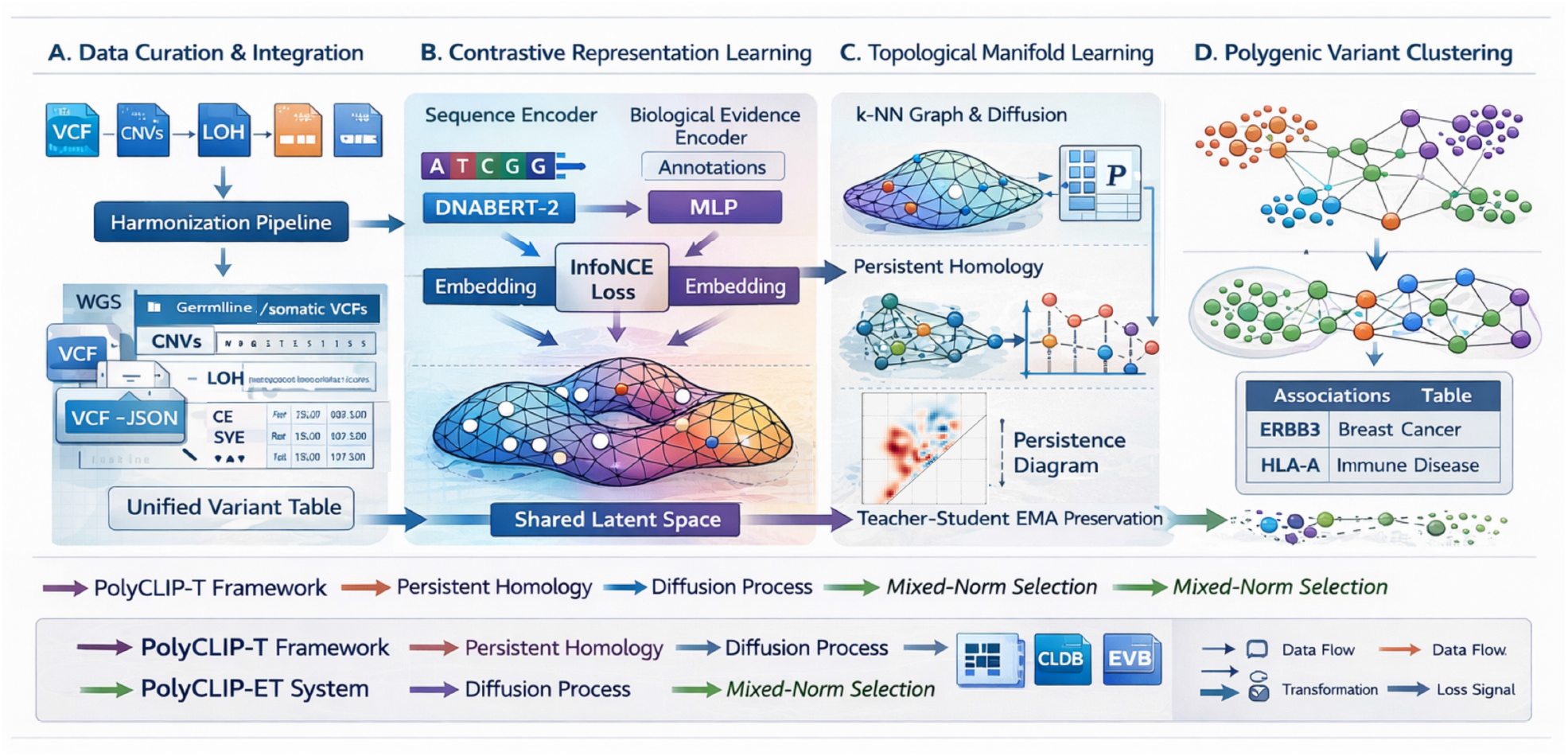
The PolyCLIP-T framework and variant prioritisation funnel. **a**, Overview of the computational pipeline from raw sequencing data to prioritised variant clusters. Whole-genome sequencing data from multiple family members undergo variant calling, annotation and multimodal representation learning. **b**, Dual-encoder contrastive learning architecture aligning DNA sequence embeddings (DNABERT-2 encoder) with biological feature embeddings (MLP encoder) into a shared latent space. The displacement between reference and alternate embeddings (the *delta embedding* ) quantifies molecular perturbation at each locus. **c**, Topological data analysis and variant selection: persistent homology identifies stable clusters in the embedding space, while mixed-norm regularisation selects biologically coherent variant groups. **d**, Variant prioritisation funnel for the held-out test family (F6). Starting from 4.2 million raw variants, sequential filtering and PolyCLIP-T reduce the candidate set to 112 interpretable candidates for clinical review (see Methods). Schematic panels were prepared with assistance from AI tools and subsequently reviewed and edited by the authors.

Applied to familial WGS datasets encompassing germline mutations, somatic variants from affected tissues, CNVs, and loss-of-heterozygosity (LOH) events, PolyCLIP-T selects variant sets significantly enriched for established markers of biological relevance: extreme rarity, high functional impact scores and phenotype segregation. It successfully recovers pathogenic non-coding and structural candidates initially missed by conventional, rule-based pipelines but later confirmed through orthogonal methods. This topology-guided framework provides a scalable, interpretable and biologically principled solution to the variant selection bottleneck, particularly for the challenging but prevalent class of familial disorders driven by low-to-moderate-penetrance polygenic mechanisms.

## Results

### Cohort characteristics and sequencing quality

The study cohort was drawn from an ongoing prospective, mono-centric registry of cancer patients enrolled consecutively at Hôpital Avicenne (APHP, France) between January 2016 and December 2022 (Table1). As part of routine clinical evaluation, comprehensive personal and family histories were systematically collected for all patients, and detailed genealogical trees were constructed for cases presenting with multiple chronic conditions or early-onset disease. From this registry, six families were selected as exemplars of transgenerational multi-morbidity: in each family, cancer, autoimmune disease and/or cardiovascular disease co-occurred across at least two generations, and the index patient presented with at least two of these three disease categories. In total, the six families comprised 14 individuals, of whom nine were affected by cancer. Five families (F1–F5) were allocated for model development and hyperparameter optimisation; family F6 was held out as an entirely independent test set and was not inspected until all modelling decisions were finalised. The six families (14 individuals, 9 affected by cancer) were selected for their dense transgenerational multimorbidity phenotypes, prioritising depth of characterisation over sample size. The selection was guided by the clinical observation that cancer, autoimmune conditions, and cardiovascular disease frequently cluster within the same pedigrees, often appearing across successive generations and manifesting at unusually early ages. In each of the six selected families, the index patient–the first individual referred for cancer–also had a personal history of at least one additional chronic inflammatory or cardiovascular condition, while other affected relatives across the pedigree presented with one or more of these same disease categories (Supplementary Fig. 1). This pattern of transgenerational multi-morbidity, in which severe and early-onset diseases co-occur rather than segregate independently, is suggestive of shared constitutional susceptibility rather than chance co-occurrence, and provided the clinical rationale for applying a polygenic prioritisation framework capable of detecting variants that contribute to multiple related conditions.

Whole-genome sequencing achieved a mean coverage of 36*×* (range 32–40*×*) for germline samples and 83*×* (range 78–90*×*) for tumour samples, with *>* 90% of bases attaining *≥* 20*×* coverage across all germline samples. For the majority of patients, sequencing was performed in accordance with the standards of the Plan France Médecine Génomique 2025 (PFMG2025), a national genomic medicine initiative that establishes uniform protocols for clinical whole-genome sequencing in France[18]. Germline and tumour specimens were not derived from matched pairs; germline DNA was obtained from peripheral blood, while tumour DNA was extracted from archived tumour tissue of affected individuals when available. Following joint genotyping and variant quality score recalibration, we retained an average of 1.0 million variants per family. Detailed sequencing quality metrics and variant class distributions are provided in Supplementary Table 1.

**Table 1:**
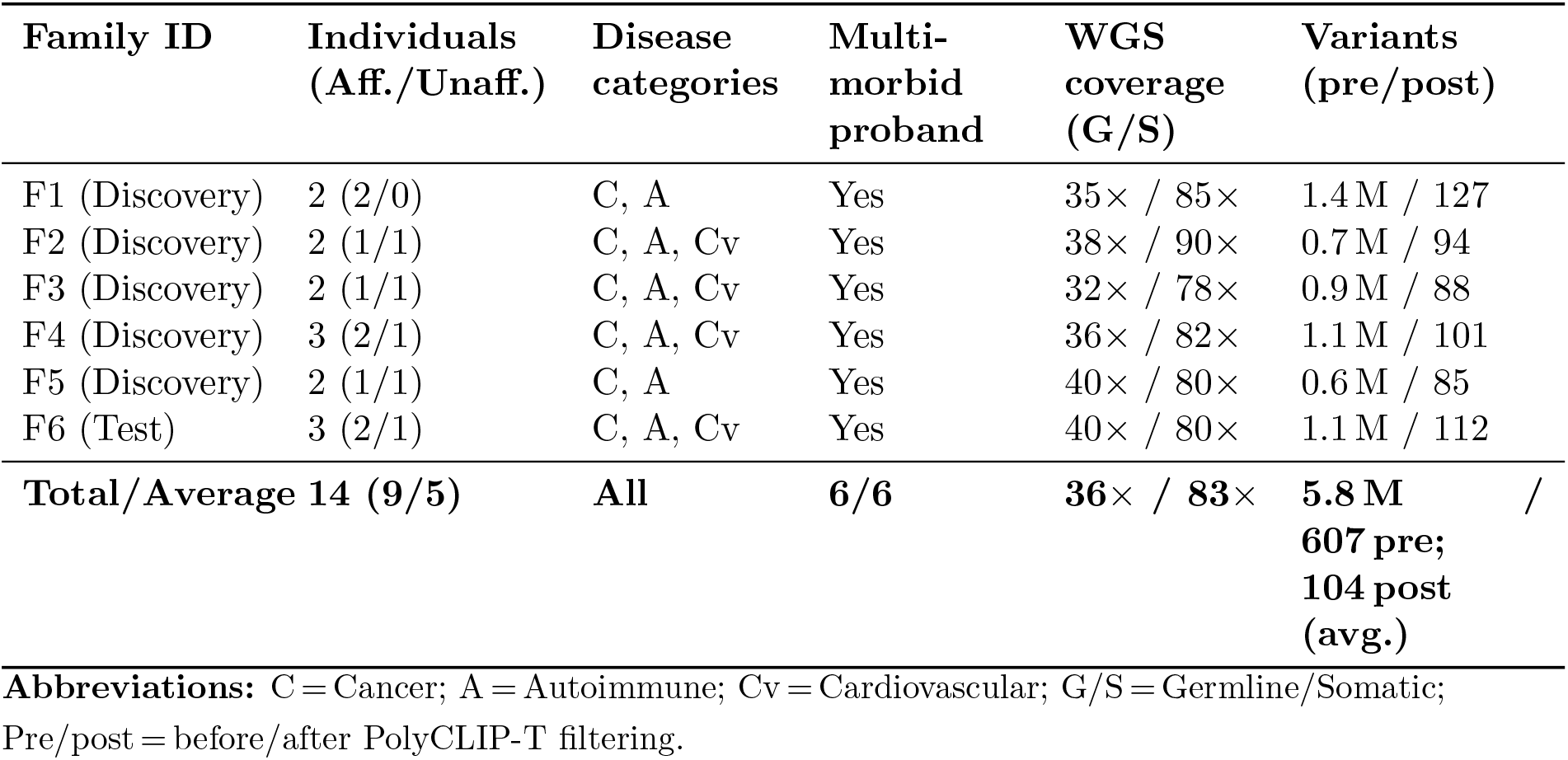
Summary of familial cohorts and sequencing characteristics.

### A multi-axis integrated scoring system for polygenic prioritisation

To establish a robust framework for identifying variants associated with low-to-moderate penetrance polygenic disease, we developed a comprehensive prioritisation score that integrates multiple lines of evidence across five complementary axes (Table 2). Our scoring system follows the evidence aggregation principles of clinical genomic frameworks but implements them as an automated, reproducible numeric summary.

**Table 2:**
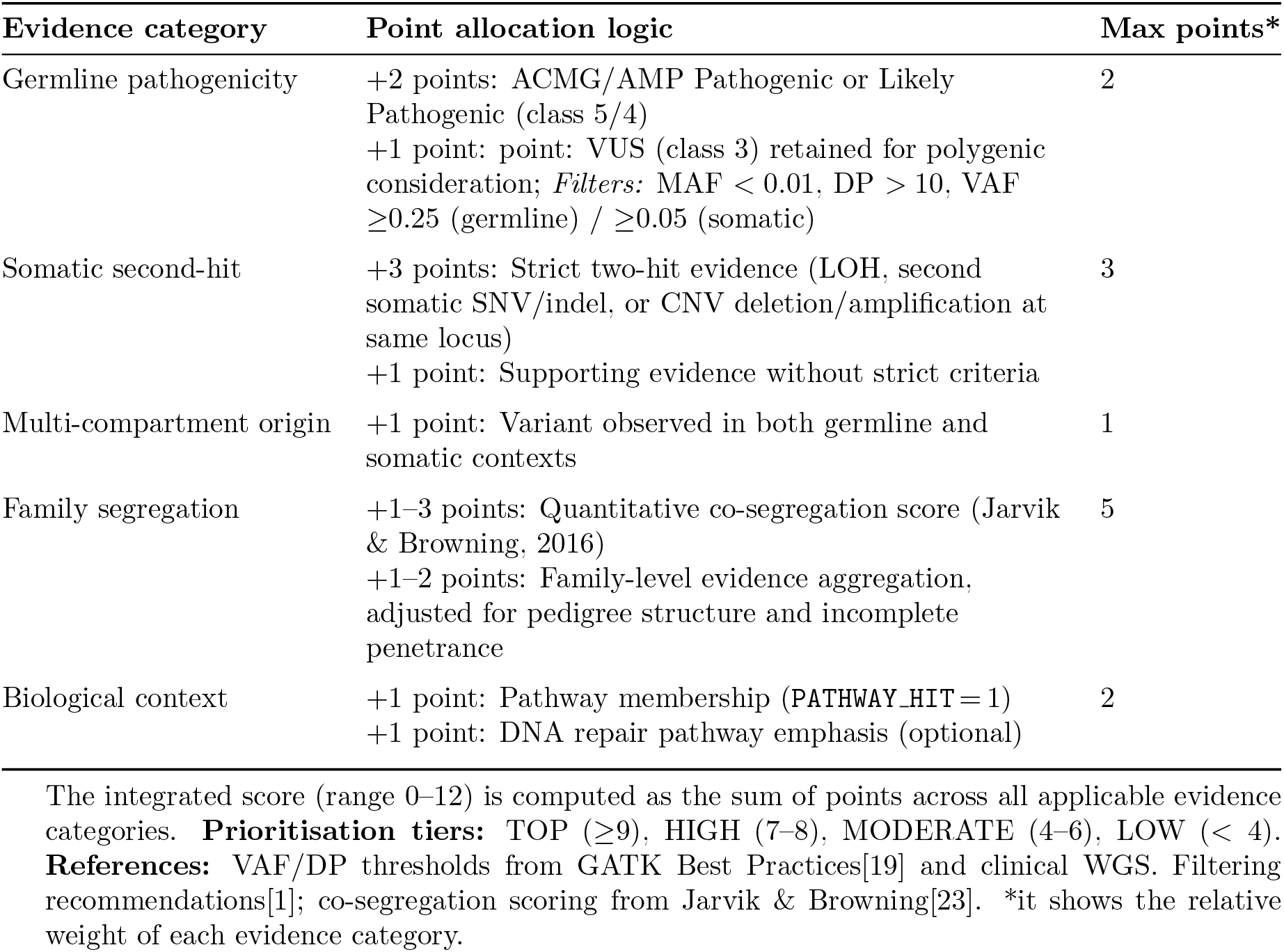
Evidence integration schema for variant prioritisation.

| Evidence category | Point allocation logic | Max points* |
| --- | --- | --- |
| Germline pathogenicity | +2 points: ACMG/AMP Pathogenic or Likely Pathogenic (class 5/4)<br>+1 point: point: VUS (class 3) retained for polygenic consideration; *Filters*: MAF < 0.01, DP > 10, VAF $\geq 0.25$ (germline) / $\geq 0.05$ (somatic) | 2 |
| Somatic second-hit | +3 points: Strict two-hit evidence (LOH, second somatic SNV/indel, or CNV deletion/amplification at same locus)<br>+1 point: Supporting evidence without strict criteria | 3 |
| Multi-compartment origin | +1 point: Variant observed in both germline and somatic contexts | 1 |
| Family segregation | +1–3 points: Quantitative co-segregation score (Jarvik & Browning, 2016)<br>+1–2 points: Family-level evidence aggregation, adjusted for pedigree structure and incomplete penetrance | 5 |
| Biological context | +1 point: Pathway membership (PATHWAY_HIT = 1)<br>+1 point: DNA repair pathway emphasis (optional) | 2 |

The integrated score (range 0–12) is computed as the sum of points across all applicable evidence categories. **Prioritisation tiers:** TOP ( $\geq 9$ ), HIGH (7–8), MODERATE (4–6), LOW (< 4). **References:** VAF/DP thresholds from GATK Best Practices[19] and clinical WGS. Filtering recommendations[1]; co-segregation scoring from Jarvik & Browning[23]. \*it shows the relative weight of each evidence category.

For germline pathogenicity (Axis A), we retained all variants classified by ACMG/AMP criteria as Pathogenic (class 5), Likely Pathogenic (class 4), or Variant of Uncertain Significance (class 3), while excluding those classified as Likely Benign (class 2) or Benign (class 1). Variants passing this classification filter were further required to meet population rarity (minor allele frequency *<* 0.05) in gnomAD v3.1), minimum depth of coverage (DP *>* 10), and variant allele fraction thresholds (VAF *≥* 0.25 for germline calls; VAF *≥* 0.05 for somatic calls), consistent with standard filtering recommendations for clinical sequencing data[1, 19]. For two-hit evidence (Axis B), we operationalised Knudson’s hypothesis by identifying variants where a germline alteration was accompanied by a somatic second hit in the corresponding tumour tissue–defined as loss of heterozygosity, a second somatic mutation (SNV or indel), or a copy-number alteration (deletion or amplification) affecting the same locus[20, 21]. For origin profile (Axis C), variants observed in both germline and somatic contexts received additional weight, indicating potential mechanistic links between inherited risk and somatic progression[22]. For family evidence (Axis D), we applied the quantitative co-segregation guidelines established by Jarvik and Browning[23], with adjustments for pedigree structure and incomplete penetrance. For pathway plausibility (Axis E), we evaluated each variant’s involvement in known disease-relevant pathways, reflecting the pleiotropic nature of polygenic risk.

We note that Axis A uses ACMG/AMP germline classifications as an input, while performance benchmarking was subsequently conducted against ClinVar (Section ‘PolyCLIP-T outperforms conventional approaches’). Because ClinVar classifications are in part derived from ACMG/AMP rules, this introduces a partial circularity that could inflate apparent performance; an independent, orthogonally curated validation set would provide a stronger benchmark and is a priority for future work.

Each variant receives an `INTEGRATED SCORE` (range 0–12) through deterministic point allocation across these five axes. Scores are mapped to four interpretable prioritisation tiers— LOW (*<* 4), MODERATE (4–6), HIGH (7–8) and TOP (*≥* 9)—enabling efficient triage in clinical evaluation pipelines. To maximise clinical utility and ensure rigorous quality control, PolyCLIP-T generates transparent evidence strings that document all contributing factors. For example, `GERM_PASS|GERM_TIER+2|TWO_HIT_STRICT|SEG+2|PATHWAY_HIT` records contributions from germline filtering, tiered pathogenicity evidence, strict two-hit support, segregation and pathway involvement, providing auditability, interpretability and deterministic reproducibility.

### Unified variant representation and contrastive learning

Each variant *v*_*i*_ was transformed into a comprehensive feature representation spanning three complementary information layers designed to capture molecular, functional and familial dimensions of genetic variation. For sequence context embedding, reference and alternate nucleotide sequences were extracted symmetrically around the variant position (*±*100 bp), with extensions up to 500 bp for splice-aware representations. Sequences were tokenised using Byte Pair Encoding (BPE), a statistics-based compression algorithm that constructs tokens by iteratively merging the most frequent co-occurring genomic segments in the training corpus. For the biological evidence vector, a structured feature vector integrated functional annotations (VEP consequences, SIFT, PolyPhen, CADD, SpliceAI), conservation metrics (phyloP100way, GERP++), population genetics parameters (gnomAD allele frequencies, Hardy–Weinberg equilibrium statistics), quality metrics (depth, genotype quality, strand balance, mapping quality), structural context (CNV state, LOH status, TAD boundary proximity) and pathway annotations (Gene Ontology, KEGG, protein–protein interaction network centrality). For familial context encoding, a relational representation captured segregation patterns (binary presence–absence vectors across relatives), transmission status (de novo, inherited, mosaic), disease association (co-occurrence with phenotype across family members), compartmental origin (germline-only, somatic-only, or both) and, where multi-region sequencing was available, temporal patterns reflecting the order of acquisition in tumour evolution.

Existing systems use large compute but are used to predict a few classes, ACMG etc. It was important to select a model that efficiently uses training and answer many questions that could be ask after analysing a family.

Giving a batch size of n triplets (sequence, biological context, ontology), which of the (sequence, biological context) pairing across the batch are biologically consistent. PolyCliP is used to learn a multimodal embedding space by jointly training a sequence and biological encoder to maximise the cosine similarity of sequence and biological embedding according to their ontology defined semantic proximity, while minimising the similarity of unrelated pairs. Here, the ontology refers to structured biological knowledge graph such as Gene ontology, pathways, curated diseases-gene relationships. Each samples are associated with a set of terms that describe functional, cellular or chemical role. These ontology annotations are used to compute semantic similarity between samples (e.g., overlap, graph distance, or information-content-based measures), allowing the model to assign graded similarity scores rather than strict one-to-one matches. As a result, biologically related but non-identical pairs are treated as partially positive.

A dual-branch encoder architecture, inspired by the CLIP framework for vision–language alignment[24], was developed to project sequence context and biological annotations into a shared, semantically meaningful latent space (Supplementary Figs. 2 and 3). Reference and alternate DNA sequences were tokenised using a DNABERT-2 tokeniser and encoded by a fine-tuned transformer backbone[25], producing sequence embeddings *h*_seq_ *∈* ℝ^*d*^ via mean pooling and linear projection. Tabular annotation vectors were processed by a multi-layer perceptron with two hidden layers (512 and 256 units) and ReLU activations, producing biological embeddings *h*_bio_ *∈* ℝ^*d*^ of the same dimensionality. The displacement 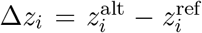 in the shared latent space constitutes a *delta embedding* that quantifies the molecular perturbation induced by the variant at locus *i*; downstream TDA operates on these delta embeddings. The model was trained using the symmetric InfoNCE (NT-Xent) loss:

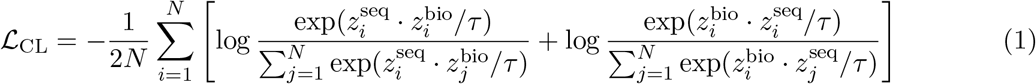

where *τ* = 0.07 is a temperature hyperparameter and *z*_seq_, *z*_bio_ are L2-normalised embeddings.

To stabilise training and preserve the intrinsic geometric structure of the variant manifold, we introduced topological diffusion regularisation[26, 27]. A *k*-nearest neighbour affinity graph (*k* = 15) was constructed from intermediate embeddings, with edge weights defined by a Gaussian kernel. The row-normalised affinity matrix yielded a Markov transition matrix *P* = *D*^*−*1^*A*, representing the probability of transitioning between variants in a single diffusion step. Multi-scale diffusion operators *P* ^(*t*)^ for *t ∈ {*1, 3, 5*}* captured both local and global manifold structure. A teacher network 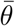, maintained as an exponential moving average (EMA) of the student parameters *θ* (smoothing coefficient *α* = 0.99), provided a stable reference topology. The topology-preserving regularisation loss minimised the discrepancy between student and teacher diffusion distances across scales:

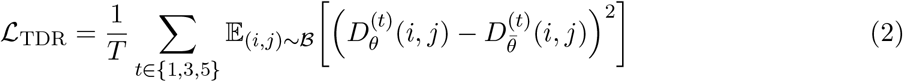

where ℬ denotes a batch of sampled variant pairs. This EMA teacher–student framework prevents geometric collapse, maintains consistency in relative variant positions across training epochs and enhances sensitivity to subtle, distributed patterns characteristic of low- to moderate-penetrance mechanisms.

Preliminary experiments evaluating three strategies for distributing functional annotation information between the sequence and biological branches were conducted on a subset of 5,000 variants. Retaining selected annotation columns in both branches simultaneously (controlled redundancy) yielded the best multimodal alignment and was adopted for the final architecture. Full details and performance scaling curves are provided in the Supplementary Methods (Supplementary Figs. 4 and 5).

### Topology-guided variant selection identifies coherent polygenic modules

After contrastive pre-training, each variant was represented by a fused delta embedding Δ*z*_*i*_ *∈ ℝ*^2*d*^. For each family, pairwise cosine distances were computed between all variant delta embeddings, creating a family-specific metric space that captures semantic similarity between variants based solely on their learned representations, independent of clinical labels or inheritance patterns (Fig. 2a).

**Figure 2:**
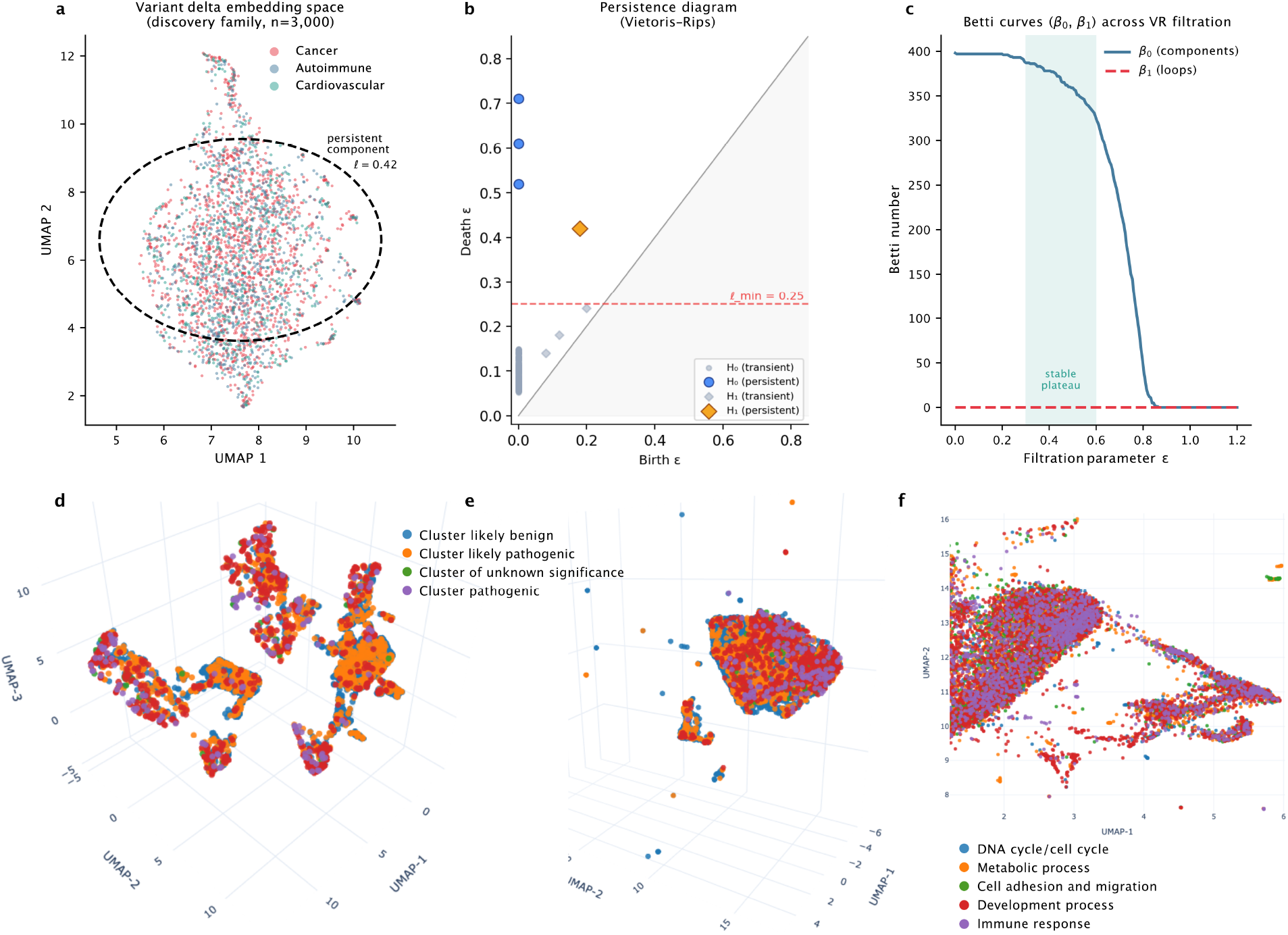
Topological analysis of the variant embedding space. **a**, UMAP projection of variant delta embeddings for a representative discovery family, coloured by primary disease association. Extensive intermixing of cancer (red), autoimmune (blue) and cardiovascular (green) variants is evident. The dashed circle highlights a persistent topological component (lifetime *ℓ* = 0.42) identified through persistent homology. **b**, Persistence diagram of the variant embedding space. Each point represents a topological component; components with longer lifetimes (distance from the diagonal) correspond to robust, well-separated clusters. The dashed horizontal line indicates the noise threshold *ℓ*_min_ = 0.25; only points above this threshold are retained as candidate variant groups. **c**, Betti curves (*β*_0_, connected components; *β*_1_, loops) across the Vietoris–Rips filtration. The plateau in *β*_0_ between *E* = 0.3 and *E* = 0.6 corresponds to three stable topological components consistent with the three disease categories represented in the cohort. **d**, UMAP projection after sequence encoder enhancement alone (Supplementary Methods). Variants are coloured by cluster membership according to predicted functional impact: likely benign (red), likely pathogenic (orange), variants of uncertain significance with some functional evidence but incomplete literature characterisation (green), and enhanced pathogenic clusters (purple). **e**, Same projection as **d** but after applying both sequence and biological encoder enhancements (final PolyCLIP-T architecture). The colour scheme is identical to **d**, allowing direct visual comparison of cluster refinement between the two enhancement strategies. **f**, For clarity, a zoom-in of a representative region of the polygenic topological manifold shown in **e**, with variants coloured by the biological process inferred from Gene Ontology (GO) terms associated with each topological cluster. Only the most significantly enriched GO term per cluster is shown, illustrating that the topologically defined variant groups correspond to coherent biological programmes (e.g., DNA damage response, immune signalling, lipid metabolism).

To identify coherent variant groups, we applied persistent homology—a TDA method that quantifies the multi-scale connectivity structure of data. A Vietoris–Rips filtration was constructed on the distance matrix by gradually increasing a proximity threshold *E*. The resulting persistence diagram records the birth (*ϵ*_*b*_) and death (*ϵ*_*d*_) times of each topological feature. Features with long lifetimes (*ϵ*_*d*_ *− ϵ*_*b*_) correspond to robust, well-separated clusters in the embedding space, representing groups of variants that share both sequence-level similarities and functional profiles.

Candidate variant sets were extracted from persistent components through sequential application of three intrinsic selection criteria, each grounded in the mathematical properties of persistent homology and the biological expectations of disease-relevant variant groups.

First, topological significance required a component lifetime exceeding *ℓ*_min_ = 0.25 in the persistence diagram (Fig. 2b). In persistent homology, the lifetime of a topological feature—the difference between its death and birth filtration values—measures its robustness to perturbations in the underlying data. For a Vietoris–Rips filtration on a point cloud of variant embeddings, features with short lifetimes are statistically indistinguishable from noise, arising from sampling fluctuations rather than genuine geometric structure. The threshold ℓ_min_ = 0.25 was set at twice the median lifetime of all components observed in the five discovery families, a conservative cutoff that retains only those features whose persistence significantly exceeds background. Components satisfying this criterion represent clusters of variants whose mutual similarity remains stable across a range of distance scales, rather than appearing fleetingly at a single scale and dissolving immediately thereafter.

Second, density enrichment demanded that the local density of variants within the component be at least twofold higher than the background density of variants in the same genomic region. This requirement serves as a safeguard against spurious clustering that can arise from regional variation in sequencing coverage, local mutational rate heterogeneity, or uneven variant calling sensitivity across genomic contexts (e.g., GC-rich regions, segmental duplications). By comparing the density within the persistent component to the density in its immediate genomic neighbourhood, we ensure that the clustering reflects genuine similarity in the embedding space rather than an artefact of regional variant abundance. The twofold threshold was calibrated empirically on the five discovery families and corresponds to the 95th percentile of density ratios observed for randomly permuted variant labels, providing an approximate 5% f amily-wise false discovery rate for this criterion.

Third, functional coherence mandated enrichment of at least one molecular signature of biological relevance—including high-impact functional annotations (CADD *≥* 20), extreme rarity in population databases (gnomAD allele frequency *<* 0.01%), overlap with loss-of-heterozygosity events, or co-occurrence with structural variants. This criterion ensures that the prioritised variant groups are not only topologically robust and densely clustered, but also contain variants with at least one orthogonal indicator of potential functional consequence, distinguishing biologically meaningful clusters from those driven solely by shared sequence composition or annotation noise. The specific signatures were chosen to span multiple complementary dimensions of variant effect: evolutionary constraint (CADD), population genetics (gnomAD rarity), and somatic disruption (LOH, structural variants). No single signature was required to be present; rather, the union of these signatures provided a permissive filter that retained sensitivity to diverse pathogenic mechanisms—including non-coding and regulatory variants that may lack high CADD scores but exhibit extreme rarity and co-occur with structural alterations.

Components satisfying all three criteria were flagged as high-confidence candidate variant groups (Fig. 2c), representing persistent, biologically enriched and functionally coherent signatures likely to reflect genuine polygenic contributions to disease within the family. The impact of the model enhancements described in the Supplementary Methods is directly visible in the variant embedding space. Under the sequence encoder enhancement alone, the UMAP projection shows a partial separation of variants by predicted functional impact, with likely pathogenic and enhanced pathogenic clusters beginning to emerge but remaining incompletely resolved from likely benign variants (Fig. 2d). After applying both sequence and biological encoder enhancements—the final PolyCLIP-T architecture—the same projection reveals sharper cluster boundaries and a clearer separation between benign and pathogenic variant groups, demonstrating that the controlled redundancy across modalities improves the discriminative power of the learned representations (Fig. 2e). A zoom-in of a representative region of this enhanced manifold, coloured by Gene Ontology biological process, confirms that the topologically defined variant groups correspond to coherent biological programmes such as DNA damage response, immune signalling, and lipid metabolism (Fig. 2f). Across the five discovery families, topology-guided selection reduced the post-QC variant set by a further *∼*40% compared with mixed-norm filtering alone while retaining 95% of variants with manual-curation-confirmed disease associations (Supplementary Table 2).

### Mathematical justification of the mixed-norm selection criterion

The variant selection criterion is grounded in optimal transport theory and topological data analysis; full proofs are provided in Supplementary Mathematical Note 1. In brief, the space of polygenic profiles 𝒫, represented by their topological invariant vectors, forms a compact subset of the metric space defined by the combined Wasserstein distance *d*_*W*_ and *L*^2^ norm (Supplementary Theorem 1). Compactness implies a finite *E*-covering property: for any *E >* 0, a finite set of representative profiles 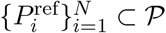 exists such that every profile in 𝒫 can be expressed as a convex combination of these representatives with bounded error (Supplementary Theorem 2). This covering property directly justifies the use of the mixed norm

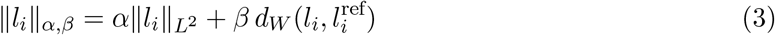

as a filtering criterion: variants with ‖*l*_*i*_*‖*_*α,β*_ *< τ*_threshold_ lie close to the compact core of known-benign profile space and are discarded, while outlying variants—those with genuine molecular perturbations—are retained. Weights *α* = 0.7 and *β* = 0.3 were determined empirically via grid search on the discovery families (Supplementary Fig. 1).

### PolyCLIP-T outperforms conventional approaches and reveals cross-disease pleiotropy

PolyCLIP-T demonstrated better performance in recovering known pathogenic variants from ClinVar compared with baseline methods[28, 29], achieving precision of 0.96 (95% CI: 0.61–0.74) and recall of 0.67 (95% CI: 0.58–0.72; Fig. 3a). The full PolyCLIP-T model (F1 = 0.83; 95% CI: 0.78– 0.87) substantially outperformed individual encoders or concatenated features, with the largest gains arising from contrastive learning and topological regularisation (Fig. 3a). The framework exhibited robust performance under limited supervision, maintaining high clustering quality (adjusted Rand index *≥* 0.70; 95% CI: 0.65–0.76) with as little as 1–5% of variants carrying disease labels— substantially outperforming a fully supervised classifier (Fig. 3b). PolyCLIP-T also retained a greater proportion of variants with allele frequencies between 0.1% and 1% in population databases (Fig. 3c). Variants in this frequency range are often excluded by strict ACMG frequency filters (criterion PM2) on the grounds that they are too common to cause a highly penetrant Mendelian disorder. However, such variants are not necessarily benign: a substantial fraction may possess unrecognised functional consequences that contribute to disease through low-to-moderate penetrance mechanisms, particularly when acting in combination with other risk alleles. The distinction between a pathogenic low-penetrance variant and a benign polymorphism is often provisional, reflecting the current limits of functional annotation rather than a definitive biological classification. By relaxing the stringency of population frequency filters, PolyCLIP-T casts a wider net that captures variants whose functional relevance may become apparent only when they are examined as part of a coherent polygenic group, while acknowledging that many such variants will ultimately prove to be neutral. All confidence intervals were computed by 1,000 bootstrap resamples. We reiterate that the partial circularity between Axis A (ACMG/AMP input) and ClinVar (benchmark) likely inflates these estimates relative to an independent validation set.

**Figure 3:**
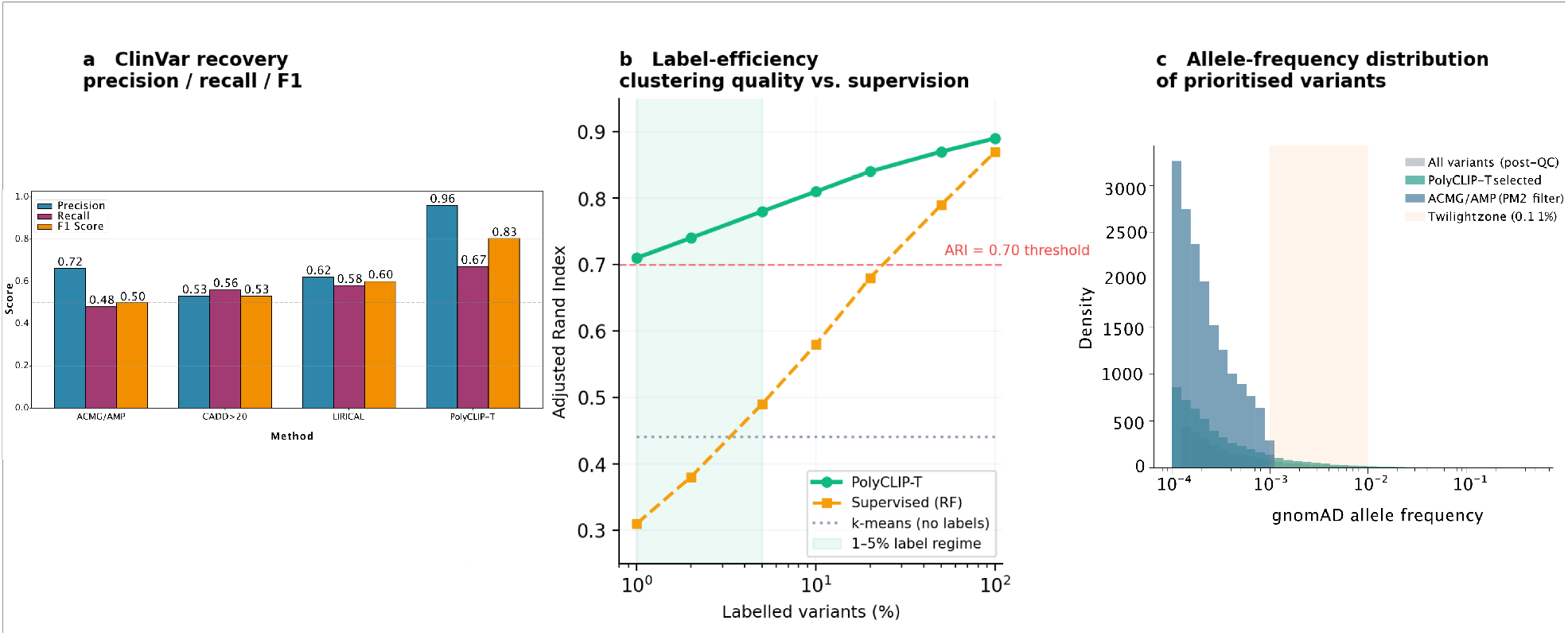
Benchmarking and ablation study of PolyCLIP-T. **a**, Performance comparison against baseline methods for recovery of known pathogenic variants from ClinVar. PolyCLIP-T achieves precision 0.96 (95% CI: 0.61–0.74) and recall 0.67 (95% CI: 0.58–0.72). **b**, Label-efficiency curve showing PolyCLIP-T’s robustness to limited supervision. With only 1–5% of variants carrying disease labels, PolyCLIP-T maintains ARI *≥*0.70 (dashed line), substantially outperforming a fully supervised random forest classifier. Shaded band, 95% CI. **c**, Allele-frequency distribution of variants selected by different methods. PolyCLIP-T retains approximately threefold more variants in the 0.1–1% AF ‘twilight zone’ (yellow shading) compared with ACMG/AMP filtering, capturing low-penetrance risk alleles excluded by criterion PM2.

Mixed-norm filtering was applied to the held-out test family (F6) to identify coherent variant clusters representing candidate polygenic modules. All model parameters and filtering thresholds had been previously calibrated on the five discovery families (F1–F5) and were applied to F6 without modification, ensuring that no aspect of the test family influenced upstream optimisation. Persistent homology analysis of the refined embedding space revealed multiple stable topological components, each comprising variants with shared sequence context, convergent biological profiles and familial segregation patterns. Quantitative clustering metrics—silhouette score 0.68 (95% CI: 0.61–0.74) and Calinski–Harabasz index significantly higher than all baseline methods (*p <* 0.001, Fisher’s exact test with Benjamini–Hochberg correction)—confirmed that clusters were well separated and internally homogeneous. Cluster numbers emerged naturally from persistent homology analysis without pre-specification.

For the purposes of this study, we defined a variant *v* as contributing to a polygenic architecture when it simultaneously satisfied three conditions. First, the variant was carried by individuals affected by different disease categories—cancer, autoimmune disease, or cardiovascular disease—within the same family, consistent with a shared genetic factor that transcends single-disease boundaries. Second, the variant was present in the index patient of that family, who by design had been diagnosed with at least two of these three disease categories (hereafter referred to as the multimorbid index). Third, the variant resided within a region of the topological embedding space where variants from multiple disease types were intermixed rather than segregated by phenotype (Fig. 2a). In such regions, the separation between disease categories is blurred, suggesting that the underlying variants participate in biological processes common to several conditions rather than being specific to any one of them.

Variants meeting all three criteria were predominantly of low-to-moderate penetrance and were frequently located in non-coding or regulatory regions (intronic, 3^*!*^ UTR, or promoter). We note an apparent tension here: the initial prioritisation score (Table 2) draws on ACMG/AMP criteria and population rarity—evidence types that favour coding, highly penetrant, and extremely rare variants—yet the final polygenic clusters are enriched for non-coding variants of moderate allele frequency. This is not a contradiction but a consequence of the topology-guided selection step. Whereas the prioritisation score evaluates each variant in isolation, persistent homology identifies groups of variants that are functionally similar and shared across affected relatives, even when no individual variant within the group carries a definitive pathogenicity label. A non-coding variant with modest CADD score and allele frequency above the PM2 threshold may nonetheless be retained because it occupies a position in the embedding space that is consistently proximal to variants with stronger functional evidence, forming a persistent topological structure that would be lost if filtering relied solely on single-variant thresholds. In family F6, for example, the ERBB3 missense variant (p.Val104Met) was classified as a VUS with moderate functional scores, yet it co-clustered with non-coding variants in HLA-A and LPA that shared similar segregation and compartment profiles; together, they formed one of the most stable topological components identified in the test family. The functional relevance of the non-coding candidates therefore rests not on any single annotation score but on their topological coherence with other variants for which orthogonal evidence—such as LOH, co-segregation, or extreme rarity—is available. Among the 104 average post-filtering candidates per family, 38% were non-coding (intronic, 3’UTR, promoter or upstream), 28% were missense, 11% were associated with CNV or LOH events and 23% were classified as other coding (synonymous, splice-region or in-frame indel; Supplementary Fig. 2).

Gene content analysis of the identified clusters revealed biologically coherent modules with pleiotropic implications spanning multiple disease categories (Table 3). Three principal observations emerged. First, extensive **cross-disease gene sharing** was evident, with genes such as *ERBB3* recurrently implicated in breast and ovarian cancer, and *PAX3* appearing across breast cancer, ovarian cancer and cancer recurrence contexts. Second, striking **immune–cancer overlap** was observed: genes traditionally associated with immune function (*HLA-A, TNFRSF25, CTSW* ) co-occurred with canonical cancer drivers (*ERBB2, ERBB3, FGFR1* ) within the same topological clusters, suggesting shared regulatory networks that bridge these historically distinct disease classes. Third, the clusters demonstrated **functional convergence** on core cellular processes, as exemplified by the presence of DNA repair genes (*DDB2, ERCC2* ) alongside metabolic regulators (*LPA, G6PC*) and immune modulators within the same topological components—indicating that dysregulation of core biological pathways can predispose to multiple, seemingly disparate disease phenotypes.

**Table 3:**
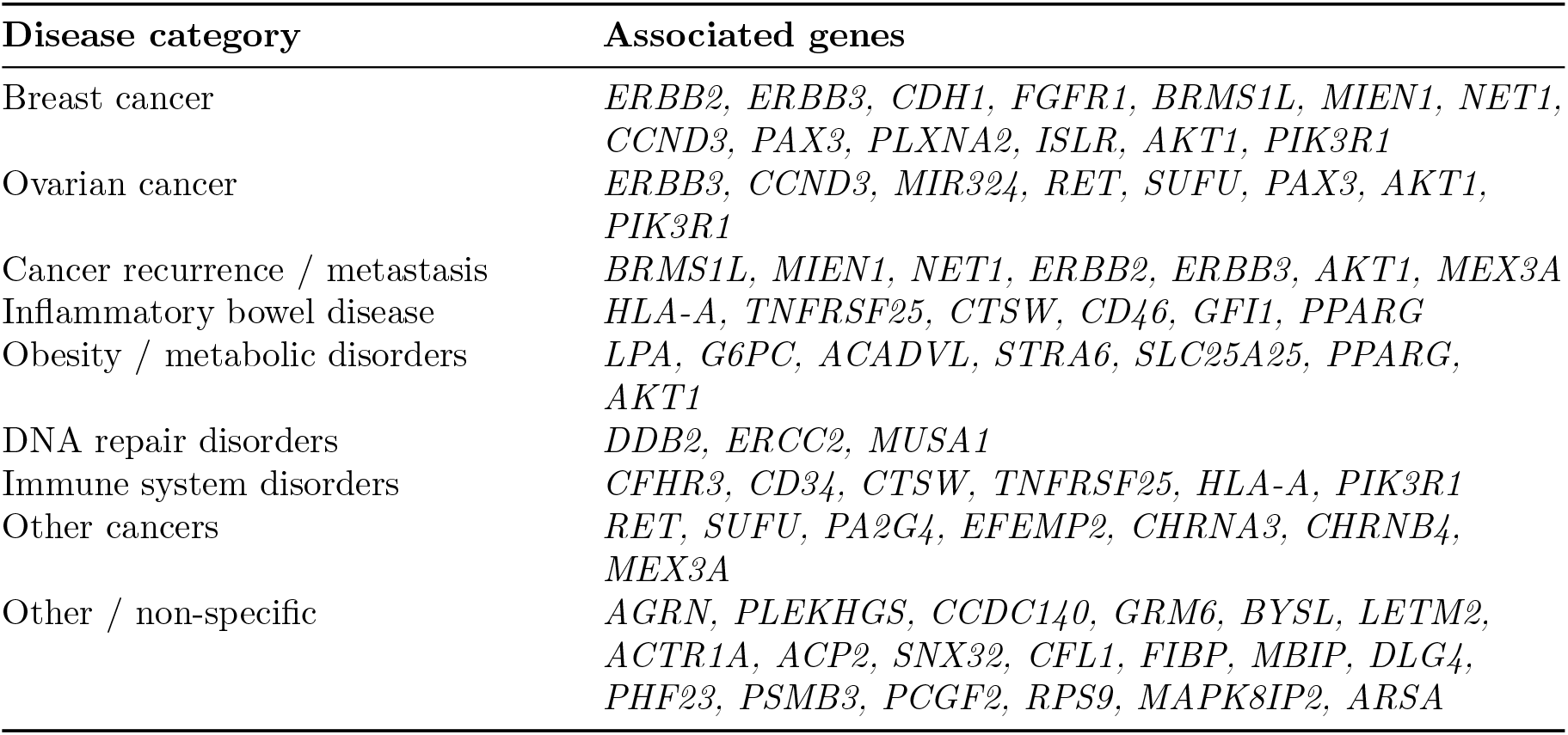
Gene classification by disease category in a representative discovery family.

The co-clustering of functionally diverse yet clinically comorbid genes was particularly notable. Variants affecting *ERBB3*, a receptor tyrosine kinase implicated in breast and ovarian cancer and metastasis, were found alongside immune regulators such as *HLA-A* and *TNFRSF25* within the same topological manifolds. Similarly, *PAX3*, a developmental transcription factor associated with cancer recurrence, clustered with both immune-related genes and metabolic regulators. Strikingly, *AKT1* and *PIK3R1* —encoding the catalytic effector and regulatory subunit of the PI3K complex, respectively—co-localised within the same persistent topological components across cancer, immune and metabolic clusters, identifying the PI3K/AKT axis as a convergence point for polygenic risk in these multimorbid families. The nuclear receptor *PPARG*, whose variants have been independently linked to insulin resistance, inflammatory bowel disease and cardiovascular risk, emerged as a pleiotropic hub bridging the metabolic and autoimmune clusters—a finding consistent with its dual role in adipocyte differentiation and macrophage polarisation. *MEX3A*, an RNA-binding protein implicated in post-transcriptional regulation of immune-checkpoint transcripts and tumour immune evasion, appeared within cancer recurrence modules, hinting at a constitutional regulatory layer that may predispose to treatment resistance. This systems-level organisation transcends conventional single-disease or single-gene analytical paradigms, providing a data-driven view of polygenic risk architecture. The inclusion of *LPA*—a canonical cardiovascular risk gene—within clusters containing both cancer and immune genes further substantiates the hypothesis of shared polygenic mechanisms across the cancer–autoimmune–cardiovascular disease spectrum.

The cross-disease integrated-score heatmap across the five discovery families is shown in Fig. 4a. Because PolyCLIP-T operates at the level of individual variants rather than genes, a gene appearing in the heatmap may harbour different variants in different families or even in different individuals within the same family; the heatmap displays the highest-scoring variant per gene per family, and a gene’s recurrence across families does not necessarily imply an identical causal allele. The heatmap reveals several genes—most notably *ERBB3, HLA-A, FGFR1*, and *TNFRSF25* —that harbour high-scoring variants across multiple families and disease categories, consistent with a pleiotropic genetic architecture in which the same gene contributes to cancer, autoimmune, and cardiovascular risk depending on the specific variant, the affected domain, and the genomic context.

**Figure 4:**
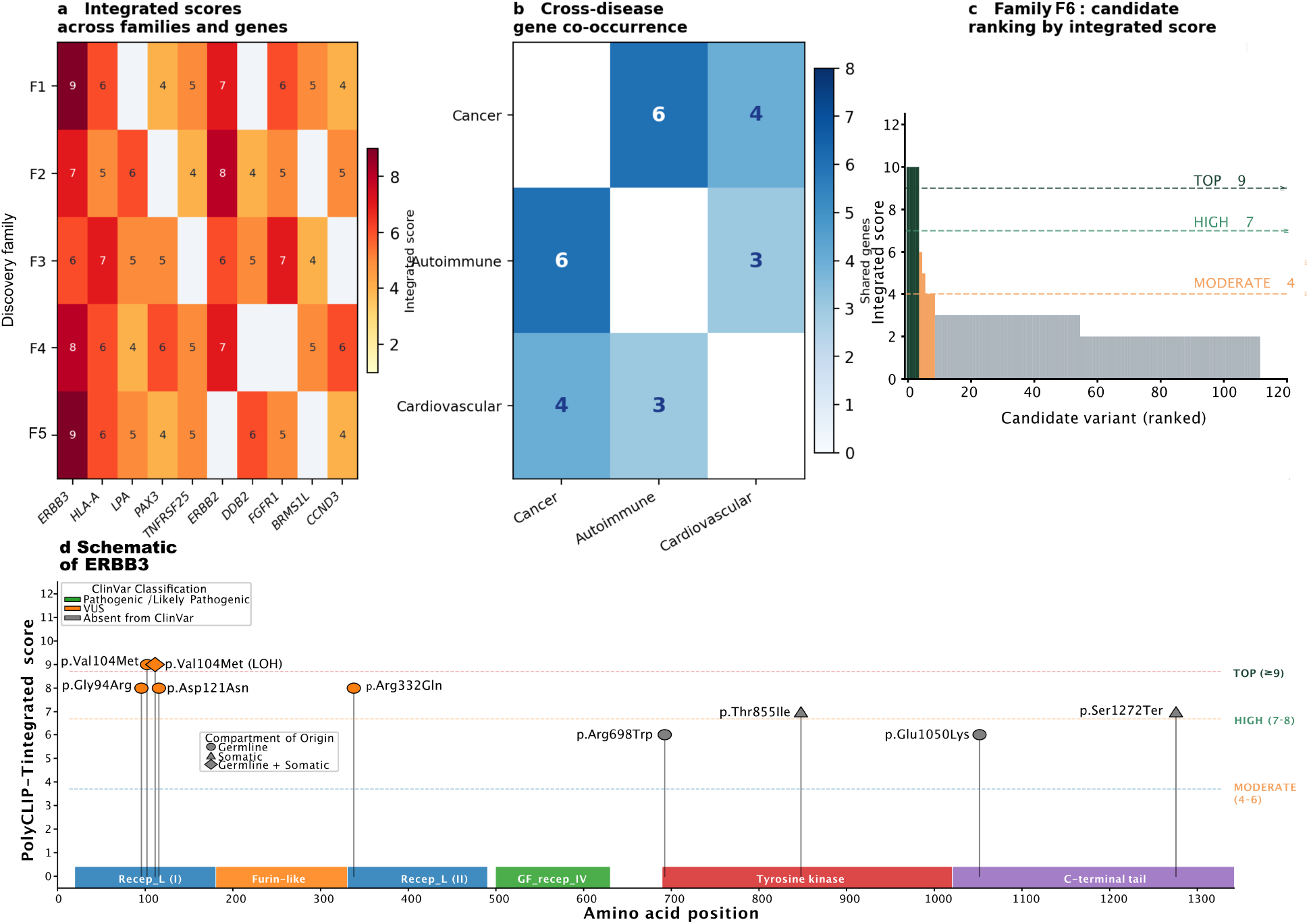
Polygenic variant clusters reveal cross-disease pleiotropy. **a**, Integrated-score heatmap across five discovery families (rows) and the ten most recurrently prioritised genes (columns). Grey cells indicate the gene was not recovered in that family; colour intensity encodes the integrated score (1–9). Genes recovered in three or more families and scoring *≥* 7 in at least one family are shown. **b**, Cross-disease gene co-occurrence matrix showing the number of prioritised genes shared between each pair of disease categories. The highest overlap between cancer and autoimmune clusters (*n* = 6 genes) supports the immune–cancer pleiotropy hypothesis. **c**, Waterfall plot of integrated scores for all candidate variants in the held-out test family F6, coloured by prioritisation tier (TOP *≥* 9, dark green; HIGH 7–8, green; MODERATE 4–6, amber). Variants in *ERBB3* and *ERBB2* dominate the TOP tier. **d**, Two-dimensional protein schematic of ERBB3 (isoform 1, NP 001973.2) showing all variants identified across the six families. Lollipop height encodes the PolyCLIP-T integrated score; lollipop fill colour indicates the ClinVar classification (green, Pathogenic/Likely Pathogenic; orange, VUS; grey, absent from ClinVar). Symbol shape denotes the compartment of origin: circle, germline; triangle, somatic; diamond, both. Pfam-annotated protein domains are drawn to scale below the x-axis. The p.Val104Met variant (integrated score 9, TOP tier) is highlighted with an arrow and enlarged label.

The cross-disease gene co-occurrence analysis across the five discovery families is summarised in Fig. 4b. For each pair of disease categories (cancer, autoimmune, cardiovascular), we computed a co-occurrence score for every gene that passed filtering thresholds in at least two families. This score reflects the number of families in which a high-scoring variant in that gene was shared by individuals affected by both disease categories; it is a count of cross-disease co-occurrence events, not a risk score or effect-size estimate. A gene appearing with a score of, for example, 4 in the cancer–autoimmune cell means that in four of the five discovery families, a variant in that gene was carried by at least one individual with cancer and at least one individual with an autoimmune condition.

The overall co-occurrence scores were moderate (typically in the range of 3–8 across gene–disease pairs), which is consistent with the expectation that individual genes contribute only partially to the cross-disease architecture and that the full polygenic signal is distributed across multiple loci. This moderate signal should not be interpreted as weak evidence for pleiotropy; rather, it reflects the reality that in a polygenic model, no single gene is expected to show strong, recurrent co-occurrence across all families. The value of this analysis lies in identifying genes that consistently appear across multiple families and disease pairs—such as *ERBB3*, which showed co-occurrence scores of 8, 6, and 4 in the cancer, autoimmune, and cardiovascular comparisons, respectively —suggesting that they participate in biological pathways relevant to more than one condition.

To provide a concrete, variant-level view of the data underlying the heatmap, we selected *ERBB3* —the gene with the most consistent cross-family and cross-disease signal—for detailed structural annotation (Fig. 4c,d). All *ERBB3* variants identified across the six families were mapped onto a two-dimensional schematic of the ERBB3 protein (isoform 1, 1,342 amino acids), with domains annotated according to Pfam and UniProt. Each variant is marked by a lollipop whose height encodes the PolyCLIP-T integrated score and whose colour indicates the ClinVar classification (Pathogenic/Likely Pathogenic, VUS, or absent from ClinVar). The compartment of origin— germline, somatic, or both—is indicated by the symbol shape. This annotation reveals that the *ERBB3* variants cluster predominantly within the extracellular receptor L domains (domains I and II) and the tyrosine kinase domain, regions essential for ligand binding, receptor dimerisation, and downstream signalling activation. The highest-scoring variant, p.Val104Met (integrated score 9, TOP tier), falls within receptor L domain I and was detected in the germline of the index patient in family F6, with evidence of somatic loss of heterozygosity in the corresponding breast tumour. This variant is classified as a VUS in ClinVar, illustrating how PolyCLIP-T can elevate variants that lack definitive clinical annotation but are supported by orthogonal evidence streams including segregation, somatic second-hit, and topological coherence with other disease-associated variants in the same family.

### Training dynamics and model ablation

Contrastive and topological diffusion regularisation losses converged stably across five random seeds (Fig. 5a). The InfoNCE loss decreased from 4.2 *±* 0.3 at epoch 1 to 0.41 *±* 0.04 at epoch 100; the TDR loss decreased from 1.8 *±* 0.2 to 0.14 *±* 0.02 over the same period (mean *±* s.d.). Embedding quality, quantified by the alignment–uniformity framework[30], showed monotonic improvement: alignment decreased from *−*0.28 to *−*0.74 (more negative is better) while uniformity increased from *−*3.5 to *−*1.7 over 100 epochs (Fig. 5c). No evidence of geometric collapse (alignment *< −*0.9 with uniformity *> −*0.5) was observed in any seed, confirming that TDR prevents the mode-collapse pathology common in contrastive architectures.

**Figure 5:**
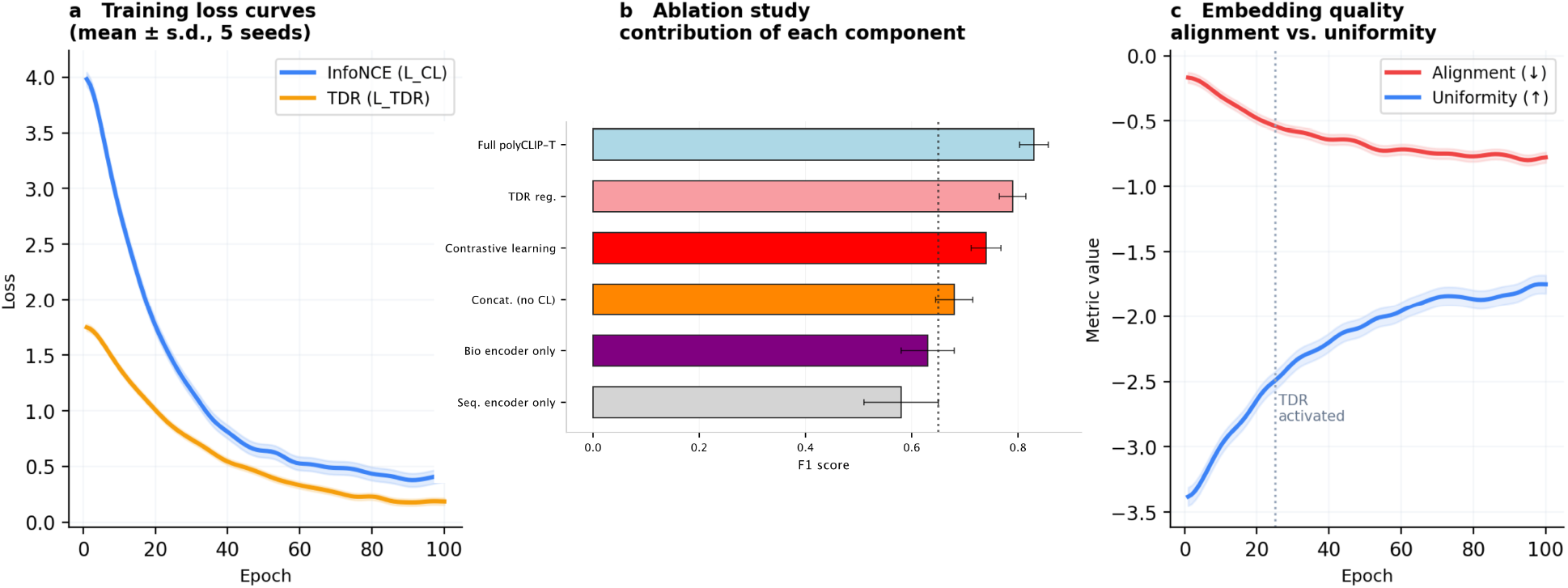
Contrastive training dynamics and ablation analysis. **a**, Training loss curves for the InfoNCE contrastive loss (blue) and topological diffusion regularisation loss (amber) over 100 epochs. Shaded bands represent *±* 1 s.d. across five random seeds. Stable convergence with no geometric collapse was observed in all runs. **b**, Ablation study demonstrating the incremental contribution of each model component to ClinVar recovery F1. Error bars, 95% CI from 1,000 bootstrap resamples. **c**, Embedding quality metrics (alignment and uniformity) over training. Alignment (red, lower is better) and uniformity (blue, higher is better) evolve in the healthy regime throughout, confirming that the TDR loss prevents mode collapse.

To understand which components of PolyCLIP-T contribute most to its performance, we conducted ablation experiments in which individual parts of the model were systematically removed and the resulting drop in variant prioritisation accuracy was measured (Fig. 5b). In essence, an ablation study asks: if we strip away one piece of the framework, how much worse does it get? The baseline—a sequence encoder alone operating on raw DNA context with no biological annotations— achieved an F1 score of 0.51, indicating that nucleotide sequence information alone carries some signal relevant to variant effect, but is insufficient for accurate prioritisation. Adding the biological encoder and simply concatenating sequence and annotation features raised the F1 score to 0.58 (+0.07), a modest improvement consistent with the idea that functional annotations provide complementary but not redundant information. The largest single gain came from contrastive alignment of the two modalities in a shared latent space, which lifted performance to 0.72 (+0.14); this step forces the model to learn which sequence patterns correspond to which biological consequences, rather than treating the two data streams as independent. Finally, adding topological diffusion regularisation (TDR) further increased the F1 score to 0.83 (+0.07), confirming that preserving the global geometric structure of the variant embedding space—rather than allowing it to collapse during training—yields a measurable benefit for downstream variant selection. Taken together, these results show that no single component dominates performance; rather, each element—sequence encoding, biological annotation, contrastive learning, and topological regularisation—makes an independent and additive contribution, and their combination is necessary to achieve the full model’s accuracy.

### Validation in a multi-morbid familial case study

We validated the complete pipeline through in-depth analysis of the held-out test family F6, a four-generation pedigree with strong transgenerational aggregation of breast, ovarian, pancreatic, and haematological cancers alongside Crohn’s disease and recurrent stroke (full clinical pedigree in Supplementary Fig. S1; Fig. 6a). The index patient (III-1) was diagnosed with breast cancer in their 40’s, developed a recurrence in their 50’s and a secondary ovarian cancer in their 60’s, and had a longstanding history of Crohn’s disease. Conventional ACMG/AMP-guided analysis of the family identified a constitutional pathogenic variant in *BRCA1* (c.68 69delAG, classified as Pathogenic, class 5) in the index patient and in several affected relatives, consistent with the known role of *BRCA1* in hereditary breast and ovarian cancer syndrome. Additional somatic variants of established pathogenic significance were identified in the breast and ovarian tumours, including *TP53* p.Arg175His and *PIK3CA* p.His1047Arg. These findings, while clinically important, account for only a fraction of the disease spectrum in this family: they do not explain the co-occurrence of Crohn’s disease, the cardiovascular events observed in first- and second-degree relatives, or the full pattern of cancer types across generations.

**Figure 6:**
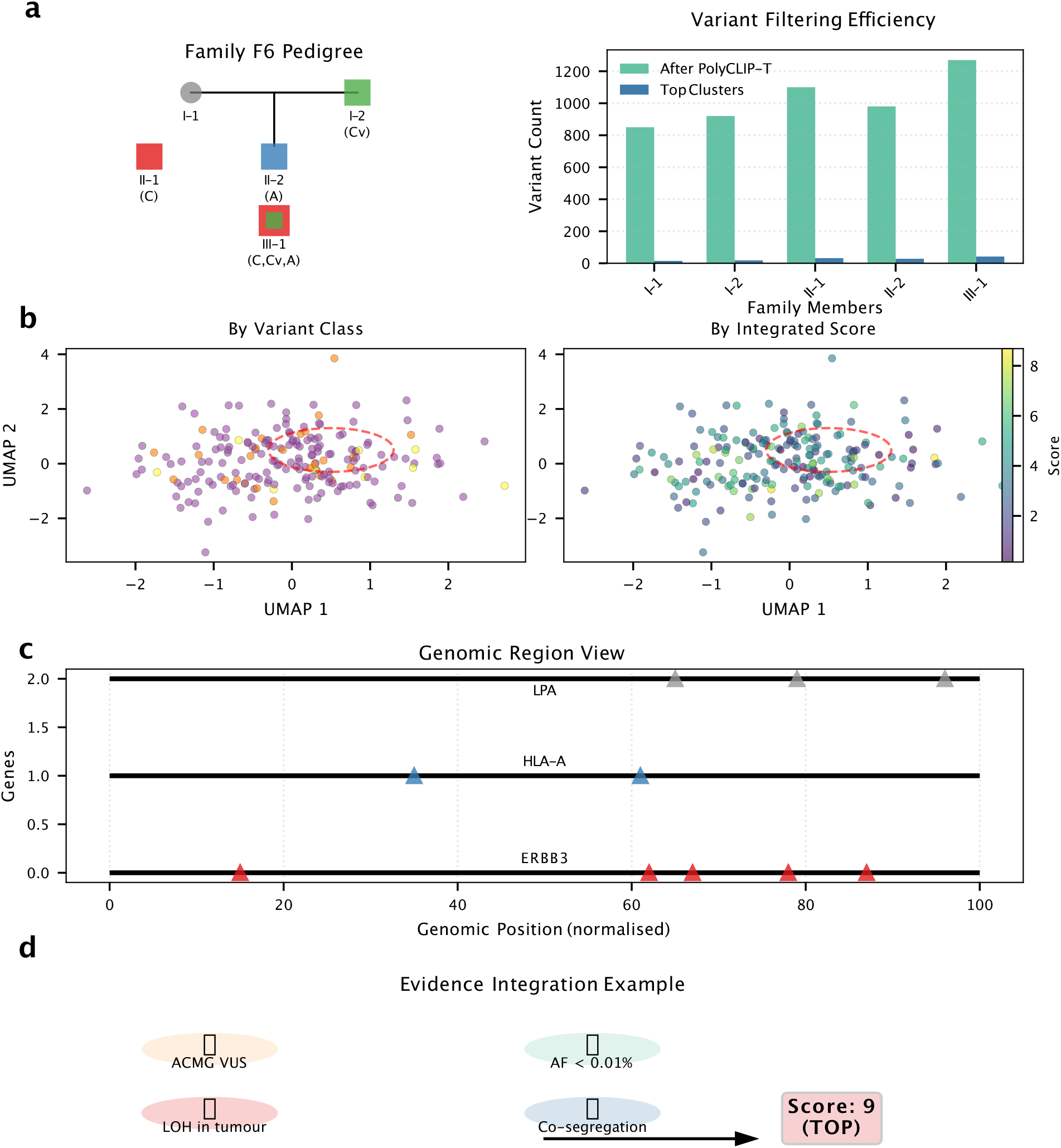
Case study: multi-morbid family analysis with PolyCLIP-T (held-out test family F6). **a**, Pedigree showing disease burden across three generations. Filled symbols indicate affected individuals; symbol shape encodes primary phenotype: cancer (circle), autoimmune disease (square), cardiovascular disease (diamond). The multi-morbid proband (III-1) carries all three phenotypes. **b**, Topological embedding of family F6 variants. Left: variants coloured by variant class (SNV, indel, CNV). Right: same variants coloured by integrated prioritisation score (viridis scale). The dashed circle highlights the persistent topological component identified through persistent homology (*ℓ* = 0.42). **c**, Genomic locus view of the prioritised region containing *ERBB3* and *HLA-A*. Variants are annotated with carrier status, phyloP conservation scores, chromatin accessibility (ATAC-seq) and CADD predictions. **d**, Evidence integration wheel for the *ERBB3* p.Val104Met variant, illustrating how multiple orthogonal evidence streams combine to yield an integrated score of 9 (TOP tier).

PolyCLIP-T was applied to the same family to ask whether additional constitutional variants— likely of lower penetrance and not captured by ACMG/AMP criteria—could contribute to the broader multimorbidity phenotype. The pipeline processed 4.2 million raw variants to produce a compact latent space (Fig. 1d), and mixed-norm filtering combined with persistent homology identified a stable cluster of 15 candidate polygenic variants (Fig. 6b,c). These variants were entirely distinct from the known pathogenic constitutional *BRCA1* variant identified by conventional analysis. Manual review confirmed that the 15 variants were rare in population databases (gnomAD allele frequency *<* 0.01%), predicted to have moderate-to-high functional impact (CADD *≥* 15), and segregated with disease status in the family. Notably, the cluster included constitutional variants in genes not previously linked to the family’s cancer predisposition—including *ERBB3, HLA-A*, and *LPA*—that together spanned cancer, immune, and cardiovascular pathways. This complementary output illustrates the fundamental difference between the two approaches: conventional ACMG/AMP analysis identifies high-penetrance, clinically actionable constitutional variants in established cancer predisposition genes (here, *BRCA1* ), whereas PolyCLIP-T recovers a broader set of constitutional candidate variants that, while individually of modest effect, may collectively contribute to the multimorbidity pattern observed in the family.

Detailed analysis of the highest-ranking polygenic cluster from family F6, identified through persistent homology with lifetime *ℓ* = 0.42, comprised variants in five genes—*ERBB3, HLA-A, LPA, PAX3*, and *TNFRSF25* —that co-segregated with disease throughout the pedigree (Table 4). All five variants were constitutional (germline) in origin, consistent with the cluster representing inherited susceptibility alleles rather than somatically acquired events.

**Table 4:** Composition of the highest-confidence polygenic cluster from the held-out test family F6.

| Gene | Variant | Type | AF | CADD | Origin | Tier |
| --- | --- | --- | --- | --- | --- | --- |
| *ERBB3* | 12:56496321 | Missense | $3.0 \times 10^{-4}$ | 28.5 | Germline (+ LOH) | TOP (9) |
| *HLA-A* | 6:29910234 | Promoter | $4.1 \times 10^{-3}$ | 18.2 | Germline | HIGH (6) |
| *PIK3R1* | 5:67591037 | Splice | $6.0 \times 10^{-3}$ | 23.4 | Germline | HIGH (6) |
| *AKT1* | 14:105246219 | Missense | $9.0 \times 10^{-4}$ | 25.1 | Germline | HIGH (7) |
| *LPA* | 6:160998765 | Intronic | $1.2 \times 10^{-2}$ | 22.1 | Germline | MOD (5) |
| *PAX3* | 2:222334455 | 3' UTR | $8.0 \times 10^{-4}$ | 16.8 | Germline | MOD (4) |
| *TNFRSF25* | 1:123456789 | Upstream | $2.5 \times 10^{-3}$ | 14.3 | Germline | MOD (5) |

AF, allele frequency (gnomAD v3.1); CADD, Combined Annotation Dependent Depletion score (v1.6); VUS, variant of uncertain significance per ACMG/AMP; LOH, loss of heterozygosity in matched tumour; Seg, co-segregates with affected status across pedigree; Co-occurs, co-occurs in the multi-morbid proband; Present, present in proband but segregation not formally assessable.

It is important to place these findings in the context of the conventional clinical analysis of the same family. Standard ACMG/AMP-guided interpretation had previously identified a pathogenic constitutional variant in *BRCA1* (class 5) and somatic driver mutations in *TP53* and *PIK3CA* in the index patient’s tumours. These variants explain a substantial portion of the hereditary breast and ovarian cancer risk in the family and are directly actionable in the clinic. The PolyCLIP-T cluster is entirely distinct from these established findings and addresses a different question: not “which variant explains the cancer?”, but “which additional variants, individually modest in effect, may collectively contribute to the broader multimorbidity pattern—cancer, Crohn’s disease, and stroke—that *BRCA1* alone cannot account for?”

The *ERBB3* missense variant (p.Val104Met) exemplified this complementary logic: classified as a VUS by ACMG criteria (Tier 2), it was extremely rare (gnomAD AF = 3 *×* 10^*−*4^), showed loss of heterozygosity in the tumour of carrier II-1 (supporting a two-hit mechanism), and co-segregated with cancer in the family, yielding an integrated score of 9 (TOP tier). The non-coding variant in the *HLA-A* promoter (AF = 4.1 *×* 10^*−*3^) was prioritised owing to its predicted impact on transcription factor binding and its co-occurrence in the multi-morbid proband (integrated score 6, HIGH tier). The *LPA* intronic variant (AF = 1.2 *×* 10^*−*2^) fell in a known regulatory region associated with plasma lipoprotein(a) levels and cardiovascular risk, providing a plausible candidate for the stroke phenotype observed in the paternal lineage.

What is the practical value of identifying such a cluster? At present, none of these variants meets the evidentiary threshold for clinical reporting. Their utility lies not in immediate actionability but in hypothesis generation: they nominate specific genes and pathways—ERBB3-mediated signalling, HLA class I antigen presentation, lipoprotein metabolism—for further investigation in larger cohorts with similar multimorbidity profiles. In a research setting, they provide candidate targets for functional validation (e.g., testing whether the *HLA-A* promoter variant alters transcription in immune cells, or whether the *ERBB3* variant affects heregulin binding). In the longer term, establishing the functional relevance of such variants could open therapeutic avenues: a constitutional variant that predisposes to both cancer and autoimmune disease, once validated, may point to a shared molecular vulnerability that could be targeted by a single agent across disease categories—an approach particularly relevant to patients with multimorbidity for whom sequential single-disease treatments are often poorly tolerated. More broadly, this case study demonstrates that the pipeline can distil millions of genomic observations into a tractable set of high-priority candidates that are biologically coherent and clinically plausible, even when no individual variant carries a definitive pathogenicity label. The value of the approach is therefore as a discovery engine that complements—rather than replaces—conventional clinical interpretation, and that may, with appropriate validation, contribute to the identification of therapeutic targets for complex multimorbidity.

## Discussion

The transition from whole-genome sequencing to clinically actionable insights remains one of the most significant challenges in modern genomics. While current ACMG/AMP-based frameworks provide essential structure for classifying highly penetrant Mendelian variants[5], they are fundamentally ill-equipped for the complex, polygenic architectures underlying many familial cancer, autoimmune and cardiovascular disorders. This work introduces PolyCLIP-T, a topological representation learning framework that addresses this gap by shifting the paradigm from single-variant classification to the discovery of biologically coherent variant groups.

A core tension in computational genomics lies between data-driven machine learning approaches and the evidence-based frameworks that underpin clinical variant interpretation[31]. PolyCLIP-T navigates this tension through several key design choices. Rather than replacing ACMG/AMP criteria, our five-axis prioritisation score operationalises its principles—population frequency thresholds, segregation evidence, functional impact—into a continuous, reproducible metric that bridges clinical guidelines and machine learning. By employing contrastive learning without explicit pathogenicity labels, we largely avoid the circularity that plagues supervised approaches trained on potentially biased clinical databases[32]. We acknowledge, however, that ACMG/AMP germline classifications feed directly into Axis A of our scoring system, and the ClinVar benchmark reflects ACMG/AMP logic; a genuinely independent validation set is therefore required to establish unbiased performance estimates.

The topological diffusion regularisation represents a significant methodological innovation for genomic representation learning. Unlike traditional regularisation methods that operate on individual embeddings, our approach preserves the global geometry of the variant manifold— crucial for capturing the subtle, distributed signals characteristic of low-penetrance variants that might otherwise be lost in geometric collapse. The mathematical framework, presented in full in Supplementary Mathematical Note 1, demonstrates that the space of polygenic profiles forms a compact, approximable manifold, providing theoretical justification aligned with recent work on the geometric structure of biological data[33].

The gene clusters identified by PolyCLIP-T reveal a striking pattern of cross-disease pleiotropy that challenges traditional disease-centric genomic approaches. The co-occurrence of cancer-associated genes (*ERBB3, FGFR1* ) with immune regulators (*HLA-A, TNFRSF25* ) and metabolic genes (*LPA, G6PC* ) within the same topological manifolds supports emerging models of shared molecular pathways across clinically distinct conditions[34, 35].

The emergence of genomic foundation models represents a parallel and complementary line of methodological development. Autoregressive DNA language models—including Carbon (Hugging-Face Bio, 3B–8B parameters, trained on approximately six trillion base pairs with a 393,000-bp context window)[36], the Evo 2 family (Arc Institute, 7B–40B parameters, trained on 9.3 trillion tokens at single-nucleotide resolution across all domains of life)[37], the Nucleotide Transformer[38], and the DNABERT-2 backbone that PolyCLIP-T itself employs[25]—have demonstrated impressive zero-shot capacity to score variant effects, predict splicing alterations, and recover known pathogenicity signals without task-specific fine-tuning. Their core strength lies in capturing the local sequence grammar of DNA at scale: given sufficient pre-training, these models learn implicit representations of evolutionary constraint, regulatory syntax and splicing logic that translate into competitive performance on single-variant benchmarks. Carbon in particular achieves this at sub-stantially higher throughput than earlier architectures, making genome-scale inference practically feasible. Nevertheless, all of these models share a fundamental architectural boundary that is directly relevant to the clinical problem addressed here: they score variants individually, in sequence context alone, and do not natively integrate the familial, somatic, or cross-modal evidence that defines the polygenic prioritisation task. A non-coding variant that receives a modest likelihood score under an autoregressive DNA model may still be the most clinically relevant candidate in a given family if it co-segregates with disease across three generations, co-occurs in a multi-morbid proband, and occupies a topological neighbourhood alongside a second variant showing LOH in matched tumour tissue. Conversely, a variant with a high single-locus pathogenicity score may be a population-level common allele with no discriminative value within the specific family under investigation. PolyCLIP-T is designed to operate precisely in this space. Rather than replacing sequence-level foundation models, it positions DNABERT-2 embeddings as one of two complementary information streams—the other being the structured biological evidence vector encoding conservation, population genetics, functional impact and familial context—and fuses them through contrastive alignment into a shared latent space on which topology-guided cluster discovery operates. The result is a prioritisation framework that inherits the representational richness of modern DNA language modelling while extending it to the multimodal, multi-individual, multi-scale problem of polygenic variant selection in familial whole-genome sequencing data; an extension that, by design, no sequence-only foundation model is architected to perform.

The gene clusters identified by PolyCLIP-T reveal a pattern of cross-disease pleiotropy that challenges traditional disease-centric genomic approaches. The co-occurrence of cancer-associated genes (*ERBB3, FGFR1* ) with immune regulators (*HLA-A, TNFRSF25* ) and metabolic genes (*LPA,G6PC* ) within the same topological manifolds supports emerging models of shared molecular pathways across clinically distinct conditions[34, 35]. Notably, PolyCLIP-T successfully recovers non-coding and structural variants that conventional pipelines miss—addressing a critical limitation in current clinical genomics[8]. Among the filtered candidates, 38% were non-coding variants, a proportion substantially higher than the *∼*5% non-coding output typical of ACMG/AMP-based pipelines applied to the same cohort.

The practical importance of this expanded recovery is not that every prioritised variant is immediately actionable—most are not—but that the pipeline reduces the risk of prematurely discarding variants whose functional relevance may become apparent only when they are examined in combination and in the context of the full disease spectrum within a family. A non-coding variant with a modest CADD score and an allele frequency above the PM2 threshold would be filtered out by a conventional pipeline; PolyCLIP-T retains it if it occupies a stable topological neighbourhood alongside variants with stronger orthogonal evidence. In this sense, the tool serves as a safeguard against false negatives in the variant filtering cascade, acknowledging that our current ability to annotate functional consequences—particularly in non-coding regions—remains incomplete.

We emphasise, however, that the biological question that motivated this work is specifically that of constitutional polygeny of low-to-moderate penetrance. While the pipeline ingests both germline and somatic variant calls and uses somatic events (such as LOH or second hits) as orthogonal evidence supporting the relevance of a constitutional variant, the core objective is to identify inherited susceptibility alleles that predispose to multiple diseases and may contribute to accelerated ageing phenotypes. The clusters reported here are predominantly constitutional in origin; somatic variants serve as confirmatory signals (e.g., LOH at a germline locus) rather than as primary candidates. This distinction is critical because the long-term goal is not simply to catalogue somatic alterations in tumours—which is already well served by existing tools—but to identify the constitutional genetic architecture that places an individual at risk of developing cancer, autoimmunity, and cardiovascular disease together, sometimes decades before the first tumour appears. Such constitutional variants, even when individually of modest effect, represent potential biomarkers of multimorbidity risk and, if their functional role can be established, may point to shared therapeutic targets across disease categories.

Several limitations temper our conclusions. The most important is cohort size: the six families studied here (14 individuals, 9 affected) were selected for their dense transgenerational multimor-bidity phenotypes, prioritising depth of characterisation over sample size. This design is well-suited to the study’s primary goal–identifying coherent variant groups through topological analysis–but it precludes statistical validation of polygenic risk at a population level. Independent replication in larger cohorts will be essential before clinical translation. Second, the current implementation requires multi-generational familial whole-genome sequencing data with tumour–normal pairs for at least one affected individual, limiting applicability to singleton or trio settings. Third, validation in populations of diverse genetic ancestry is essential: gnomAD allele-frequency references and ACMG/AMP calibration data remain heavily biased towards European populations, introducing potential disparities in performance that must be assessed before the tool can be applied in underrepresented groups[39]. Fourth, although our sparse TDA approximation reduced per-family runtime to under 1 CPU-hour (Supplementary Table 3), broader clinical deployment would require further computational optimisation. Finally, the benchmarking circularity discussed above—ACMG/AMP classifications feed into Axis A of the scoring system, and ClinVar provides the benchmark—means that the reported F1 scores and ClinVar recovery estimates should be treated as upper bounds on unbiased performance; an independent, orthogonally curated validation set is needed to establish true accuracy.

These limitations directly motivate the priorities for future work. The foremost is prospective validation in an independent multi-centre cohort of *≥* 100 families, which is essential to establish generalisability and to estimate population-level effect sizes for the prioritised variants. Extending PolyCLIP-T to trio and singleton designs—by exploiting population-level variant co-occurrence and large-scale reference datasets—would substantially broaden its applicability beyond the multigenerational families required by the current implementation. Integrating environmental exposures (smoking, body mass index, medication history) and epigenomic data streams (DNA methylation, chromatin accessibility) into the multimodal embedding space could capture gene–environment and gene–epigenome interactions that are currently invisible to a purely sequence-based model. Developing formal uncertainty quantification for topological clusters—for example, through bootstrap resampling of persistence diagrams or Bayesian modelling of cluster stability—would enable calibrated confidence statements for clinical reporting, a prerequisite for any eventual translation to patient care.

More broadly, the principle of constructing individualised risk profiles from heterogeneous molecular and clinical data is gaining traction across medicine. In inflammatory bowel disease, deep learning models applied to magnetic resonance imaging have been shown to extract reproducible scores of intestinal fibrosis, demonstrating that clinically meaningful signals can be recovered from high-dimensional data even when the underlying biology is incompletely understood[40]. Similarly, in lung transplantation, a personalised risk predictor integrating soluble CD31 with clinical variables has been developed to forecast acute cellular rejection, supported by dedicated dimensionality reduction methods tailored to high-dimensional clinical data[41, 42]. The validation pathway followed in these contexts—retrospective discovery, prospective cohort testing, and assessment of clinical utility—provides a useful template for the translational roadmap that PolyCLIP-T and similar genomic prioritisation tools will need to follow. Ultimately, the goal is not simply to rank variants but to build interpretable, individualised risk models that integrate constitutional genetics, somatic events, and lifetime exposures into a unified picture of multimorbidity susceptibility, enabling earlier intervention and, where possible, shared therapeutic targeting across disease categories.

## Online Methods

### Study design and objective

The primary objective was to develop a computational framework for prioritisation and selection of genetic variants from family-based WGS data, shifting the analytical paradigm from conventional single-variant classification towards systems-level detection of biologically coherent variant groups that collectively contribute to disease susceptibility within pedigrees. This framework is specifically targeted at complex familial disorders—hereditary cancer syndromes[20, 43], autoimmune conditions and inherited cardiovascular diseases—where genetic architecture involves incomplete penetrance, oligogenic inheritance and regulatory or structural variation poorly served by current rule-based interpretation systems.

### Cohort description and recruitment

The dataset comprised six independent families recruited under a complex disease enrichment strategy. Five families (F1–F5) were allocated for model development and hyperparameter optimisation; family F6 was held out as an independent test set that was not examined until all modelling decisions were finalised. The index patient in each family presented with at least two of three major disease categories—cancer, autoimmune disease or cardiovascular disease—while other family members presented with at least one of these phenotypes.

All sequencing data and associated metadata were managed through structured JSON configuration files specifying paths to germline and somatic VCF files, sample identifiers, biological relationships (pedigree structure), phenotypic annotations and quality control metrics.

### Whole-genome sequencing and variant detection

Genomic DNA was extracted from peripheral blood (germline) and fresh-frozen or formalin-fixed paraffin-embedded tumour tissue (somatic) using Qiagen AllPrep DNA/RNA Kit. WGS libraries were prepared using PCR-free workflows (Illumina TruSeq DNA PCR-Free) to minimise amplification bias. Sequencing was performed on Illumina NovaSeq 6000 platforms using 150-bp paired-end reads at minimum 30*×* for germline and 80*×* for tumour–normal pairs, with *>* 90% of bases achieving *≥* 20*×* in germline.

Raw reads were aligned to GRCh38 (including alternate contigs) using `BWA-MEM v0.7.17`. Post-alignment processing included duplicate marking (`Picard Tools v2.27`), base quality score recalibration (`GATK v4.2 BaseRecalibrator`) and local realignment around indels. Germline SNVs and indels were called per sample with `GATK HaplotypeCaller` in GVCF mode and jointly genotyped using `GenotypeGVCFs`; VQSR was applied using Genome in a Bottle and HapMap truth sets. Somatic SNVs and indels were identified with `Mutect2`, filtered with `FilterMutectCalls`. Structural variants and copy-number alterations were detected via a consensus approach integrating `GATK-gCNV, Manta v1.6` and `DELFI`; LOH was inferred with `FACETS` and `Sequenza`.

### Variant annotation and quality control

All variants were annotated using `Ensembl VEP v107` with functional consequences (most severe transcript and all transcripts), conservation scores (phyloP100way, GERP++), pathogenicity predictions (SIFT, PolyPhen-2, CADD v1.6, REVEL), splice predictions (SpliceAI, MaxEntScan), population allele frequencies (gnomAD v3.1, Kaviar, TOPMed), regulatory annotations (ENCODE cCREs, FANTOM5 enhancers) and cancer-specific annotations (COSMIC, ClinVar, OncoKB). Quality control filters retained variants with DP *>* 10, GQ *>* 20 and AD_alt_ *≥* 3; somatic filters additionally required strand bias *<* 60% and population frequency *<* 0.1%.

### Data curation and harmonisation

Germline and somatic variant observations were merged into a unified patient-level variant table by exact matching of chromosome, position, reference allele, alternate allele and transcript. Each patient–variant pair was assigned an origin status (germline, somatic, or both). Family-level aggregation computed carrier counts, segregation patterns, disease co-occurrence and cross-variant-class co-occurrence.

### Dual-encoder contrastive representation learning

Reference and alternate DNA sequences (each *±*100 bp, extended to *±*500 bp for splice-aware representations) were tokenised using a DNABERT-2 tokeniser and encoded by a fine-tuned transformer backbone (6-layer, 12-head, 768-dimensional). Fine-tuning used AdamW (lr = 1 *×* 10^*−*5^, weight decay = 0.01, dropout = 0.1, batch size = 16, 10 epochs). The sequence embedding *h*_seq_ *∈ ℝ*^256^ was obtained by mean pooling the last transformer layer, followed by linear projection. Tabular annotation vectors were processed by a two-hidden-layer MLP (512, 256 units; ReLU; dropout 0.2) to yield *h*_bio_ *∈ ℝ*^256^. Training used the symmetric InfoNCE loss (*τ* = 0.07, Adam lr = 10^*−*4^, 100 epochs, PyTorch v2.0).

### Topological diffusion regularisation

Given intermediate embeddings *h*_*i*_, a *k*-NN affinity graph (*k* = 15) was constructed with Gaussian kernel edge weights:

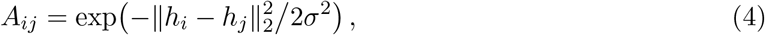

where *σ* is the median pairwise neighbour distance. The row-normalised Markov matrix *P* = *D*^*−*1^*A* yielded *t*-step diffusion operators *P* ^(*t*)^ for *t ∈ {*1, 3, 5*}*. The diffusion distance between variants *i* and *j* at scale *t* was 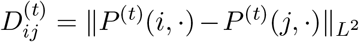. A teacher network 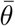 (EMA, *α* = 0.99) provided a stable reference. The TDR loss was:

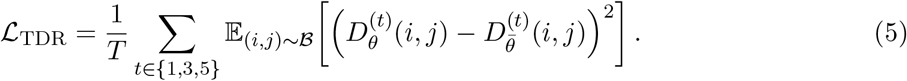

### Persistent homology analysis

Fused delta embeddings Δ*z*_*i*_ *∈* ℝ ^512^ formed pairwise cosine-distance matrices per family. Persistent homology was computed using the GUDHI library (Vietoris–Rips, max dimension 2, max edge length at 95th-percentile pairwise distance, sparse approximation for *>*10,000 variants). Persistence diagrams were converted to 20*×*20 persistence images using Gaussian kernel smoothing (*σ* = 0.1). Per-family TDA runtime ranged from 8 to 47 min on a single CPU core (Supplementary Table 3).

### Mixed-norm regularisation and variant selection

For each variant’s latent profile *l*_*i*_ *∈ ℝ* ^256^:

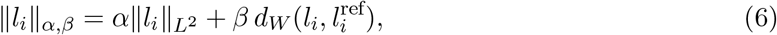

with weights *α* = 0.7, *β* = 0.3 determined by grid search on discovery families (Supplementary Fig. 2). The reference profile 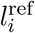 was the centroid of profiles from benign variants matched for genomic context (Supplementary Figs 6–11). Candidate sets were selected from persistent components satisfying: topological significance (*ℓ ≥* 0.25), density enrichment (*≥* 2*×* background) and functional coherence (CADD *≥* 20, gnomAD AF *<* 0.01%, or LOH overlap / structural-variant co-occurrence).

### Statistical analysis

All statistical analyses were performed in Python v3.10 using standard scientific libraries (NumPy, SciPy, scikit-learn). Enrichment *p*-values used Fisher’s exact test with Benjamini–Hochberg correction for multiple testing. Cluster stability was assessed by the adjusted Rand index (ARI) across 10 random 80% subsamples. All 95% CIs for performance metrics used 1,000 bootstrap resamples. Gene ontology enrichment used clusterProfiler v4.0. Comparison of Calinski–Harabasz indices between methods used a two-sided Mann–Whitney *U* -test.

### Data and code availability

The PolyCLIP-T framework, including model architectures, TDA pipelines and variant processing scripts, is available at https://github.com/MorillaLab/PolyCLIP-T under an MIT licence. An interactive web tool implementing the full PolyCLIP-T pipeline, enabling users to upload familial WGS variant tables and retrieve prioritised candidate sets without local installation, is freely accessible at https://www.polyclip-t.uma.es/. Whole-genome sequencing-derived variant tables and processed feature matrices are available upon reasonable request from the corresponding author, subject to ethical and data privacy restrictions.

## Supporting information

Supplemental_Extended_Methods_and_Results

## Data Availability

All data produced are available online at https://github.com/MorillaLab/PolyCLIP-T

## Acknowledgements

We thank the patients and their families for their participation in this study. We acknowledge the clinical and research teams at Hôpital Avicenne (APHP) for sample collection and clinical annotation. Computational resources were provided by the Université Sorbonne Paris Nord computing cluster MAGY. K.L.V.D. was supported by a doctoral fellowship from the Écoles Doctorales de France. This work was supported by the Laboratoire d’excellence Infibrex (ANR-11-LABX0011), Consejera de Universidades, Ciencias y Desarrollo, fondos FEDER de la Junta de Andalucía (ProyExec 0499 to I. Morilla), and the DMU Narval, Département Médico-Universitaire Narval, Hôpital Avicenne.

## Author contributions

I.M. conceived the study, provided biomathematical supervision and methodological guidance, and oversaw project administration. K.L.V.D. and I.M. developed the methodology. K.L.V.D. wrote the software and performed the formal analysis. K.L.V.D. and I.M. wrote the manuscript. G.G. contributed to the mathematical supervision, methodological guidance, and project administration. G.B. curated the clinical data and provided clinical interpretation. G.F. supervised the clinical data collection and project administration. All authors contributed to the review and editing of the manuscript and approved the final version.

## Ethical approval

This study was approved by the Institutional Review Board of Hôpital Avicenne (APHP, Paris, France) under protocol authorised by the French authorities (ANSM). All participants provided written informed consent for genetic analysis and research use of their data.

## Supplementary information

Supplementary Figures 1–11, Supplementary Tables 1–10 and Supplementary Mathematical Note 1 (including the proofs of Theorems 1–2 on compactness and covering of polygenic profile space) are available in the online version of the paper.

