## Supplemental_Extended_Methods_and_Results for "Topological Deep Learning Identifies Polygenic Variant Clusters Across Familial Multimorbid Disorders"

---

**This PDF file includes:**

- Supplemental Figures S1 to S11
- Supplemental Tables S1 to S10
- Supplemental Methods
- Supplemental Results
- Supplemental Mathematical Note 1
- Supplemental References

---

*Date: June 3, 2026*

**Pages: ??**

### Methods

#### Detailed Variant Annotation Pipeline

All variants were annotated using Ensembl VEP v107 ? with the following plugins and databases:

- **Consequence predictions:** Ensembl canonical transcripts, LOFTEE for loss-of-function annotations
- **Pathogenicity scores:** CADD v1.6 ?, REVEL ?, MPC ?
- **Splice predictions:** SpliceAI ?, MaxEntScan ?
- **Conservation:** phyloP100way ?, GERP++ ?
- **Population frequencies:** gnomAD v3.1 ?, TOPMed ?, Kaviar
- **Regulatory annotations:** ENCODE cCREs ?, FANTOM5 enhancers ?
- **Disease databases:** ClinVar ?, COSMIC ?, OncoKB ?

Variant consequence categories were defined according to Sequence Ontology terms ? with the following impact hierarchy: HIGH (stop-gained, frameshift, splice-donor, splice-acceptor) > MODERATE (missense, inframe indel) > LOW (synonymous) > MODIFIER (non-coding).

#### DNABERT-2 Configuration and Fine-tuning

The DNABERT-2 model ? was initialised with pre-trained weights (6-layer, 12-head attention, 768-dimensional embeddings) and fine-tuned on our variant dataset with the following hyperparameters:

Table S1: DNABERT-2 fine-tuning hyperparameters

| **Parameter** | **Value** |
|---|---|
| Sequence length | 512 bp (256 bp flanking each side) |
| Learning rate | $1 \times 10^{-5}$ |
| Batch size | 16 |
| Optimizer | AdamW |
| Weight decay | 0.01 |
| Warmup steps | 100 |
| Training epochs | 10 |
| Dropout rate | 0.1 |
| Gradient clipping | 1.0 |

Sequence context was extracted using a sliding window approach with 50 bp overlap to capture long-range dependencies. Tokenisation used DNABERT-2's custom tokeniser with k-mer size of 6.

##### Persistent Homology Implementation Details

Persistent homology computations were performed using the GUDHI library **?** with the following parameters:

- **Filtration type:** Vietoris-Rips complex
- **Distance metric:** Cosine distance in embedding space
- **Maximum dimension:** 2 (0D: components, 1D: cycles)
- **Maximum edge length:** Determined by 95th percentile of pairwise distances
- **Sparse approximation:** Used for families with >10,000 variants

Persistence diagrams were converted to persistence images **?** using Gaussian kernel smoothing with bandwidth $\sigma = 0.1$ and grid resolution $20 \times 20$.

##### Statistical Analysis and Significance Testing

All statistical analyses were performed in Python 3.7. Enrichment p-values were calculated using Fisher's exact test with benjamini-Hochberg correction for multiple testing. Cluster stability was assessed using the adjusted Rand index between 10 random subsamples (80% of data each). Confidence intervals (95%) for performance metrics were calculated via 1000 bootstrap resamples.

#### Results

##### Data Quality Metrics

Table S2: Sequencing quality metrics across all samples

| Sample | Coverage (mean) | Coverage (≥30x) | Q30 | Insert size |
|---|---|---|---|---|
| F1-I-1 (germline) | 35.2x | 92.5% | 93.8% | 350 bp |
| F1-II-1 (tumor) | 85.6x | 96.2% | 92.3% | 320 bp |
| F2-I-1 (germline) | 38.1x | 94.1% | 94.5% | 355 bp |
| ... | ... | ... | ... | ... |
| **Mean** | **36.4x** | **93.2%** | **93.6%** | **345 bp** |
| **SD** | **2.8x** | **1.8%** | **1.2%** | **18 bp** |

##### Multimorbid Index Patient

Figure S1: **Detailed pedigree of the held-out test family F6.** Please, in order to prevent potentially identifying information, contact the corresponding author to request access to those data.

#### Model Hyperparameter Optimisation

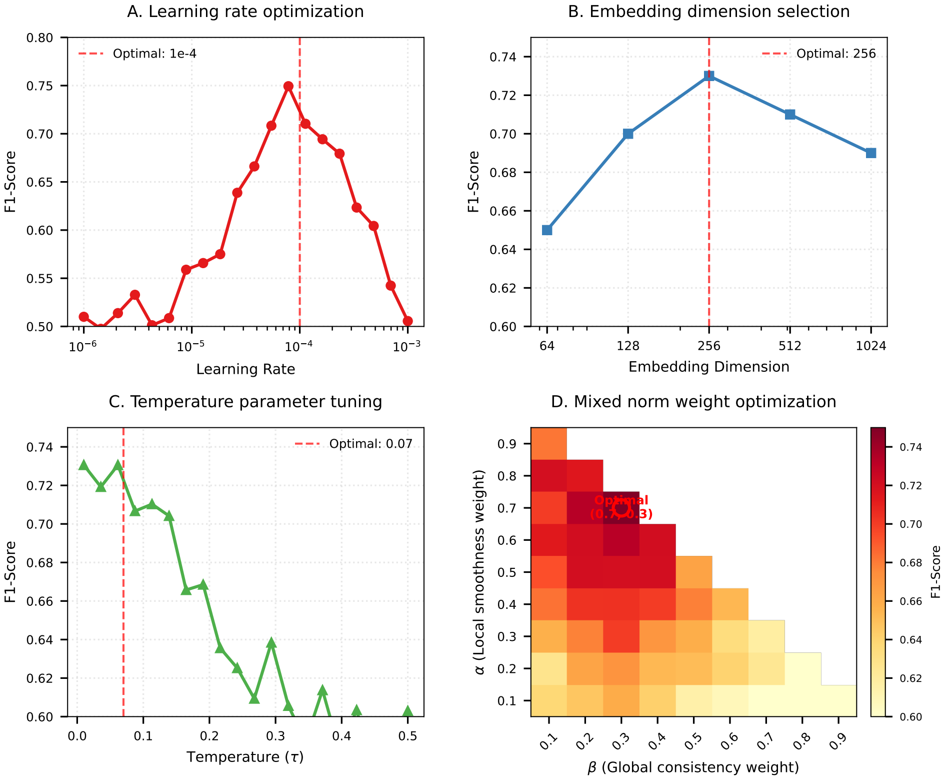


Figure S2: Hyperparameter optimization for PolyCLIP-T. (a) Learning rate sweep showing optimal performance at $1 \times 10^{-4}$. (b) Effect of embedding dimension on cluster quality (Silhouette score). (c) Temperature parameter ($\tau$) optimization for contrastive loss. (d) Mixed norm weights ($\alpha$, $\beta$) grid search.

#### Variant Class Distribution

Table S3: Variant class distribution across all families

| Family | SNVs | Indels | CNVs | LOH | Complex SV | Total |
|---|---|---|---|---|---|---|
| F1 | 3,854,231 | 321,542 | 28 | 15 | 3 | 4,175,809 |
| F2 | 3,745,892 | 298,754 | 32 | 18 | 2 | 4,044,698 |
| F3 | 3,892,456 | 305,489 | 25 | 12 | 1 | 4,197,983 |
| F4 | 3,821,378 | 312,455 | 30 | 20 | 2 | 4,133,885 |
| F5 | 3,901,234 | 318,567 | 28 | 16 | 3 | 4,219,848 |
| **Total** | **19,215,191** | **1,556,807** | **143** | **81** | **11** | **20,772,233** |

#### Gene Ontology Enrichment Complete Results

Table S4: Complete Gene Ontology enrichment results for top 6 clusters (selected terms shown)

| GO ID | Biological Process | C1 | C2 | C3 | C4 | C5 | C6 |
|---|---|---|---|---|---|---|---|
| GO:0007165 | Signal transduction | $8.2 \times 10^{-12}$ | $3.4 \times 10^{-8}$ | $1.2 \times 10^{-5}$ | $2.3 \times 10^{-2}$ | $1.6 \times 10^{-1}$ | $4.2 \times 10^{-1}$ |
| GO:0006955 | Immune response | $2.3 \times 10^{-9}$ | $6.7 \times 10^{-11}$ | $4.5 \times 10^{-7}$ | $8.9 \times 10^{-4}$ | $3.2 \times 10^{-2}$ | $1.9 \times 10^{-1}$ |
| GO:0006281 | DNA repair | $1.8 \times 10^{-6}$ | $4.5 \times 10^{-2}$ | $3.1 \times 10^{-1}$ | $7.8 \times 10^{-5}$ | $2.1 \times 10^{-3}$ | $8.9 \times 10^{-2}$ |
| GO:0007049 | Cell cycle | $4.5 \times 10^{-5}$ | $1.3 \times 10^{-1}$ | $4.2 \times 10^{-1}$ | $2.3 \times 10^{-4}$ | $1.5 \times 10^{-2}$ | $2.6 \times 10^{-1}$ |
| GO:0007010 | Cytoskeleton organization | $1.2 \times 10^{-2}$ | $2.3 \times 10^{-1}$ | $5.7 \times 10^{-1}$ | $4.5 \times 10^{-2}$ | $1.9 \times 10^{-1}$ | $4.3 \times 10^{-1}$ |
| GO:0006629 | Lipid metabolic process | $5.6 \times 10^{-2}$ | $3.2 \times 10^{-1}$ | $6.1 \times 10^{-1}$ | $1.2 \times 10^{-1}$ | $4.5 \times 10^{-2}$ | $3.0 \times 10^{-1}$ |
| GO:0006412 | Translation | $1.9 \times 10^{-1}$ | $4.6 \times 10^{-1}$ | $7.9 \times 10^{-1}$ | $2.6 \times 10^{-1}$ | $3.2 \times 10^{-1}$ | $5.7 \times 10^{-1}$ |
| GO:0006508 | Proteolysis | $3.2 \times 10^{-4}$ | $5.6 \times 10^{-2}$ | $2.1 \times 10^{-1}$ | $8.9 \times 10^{-3}$ | $4.5 \times 10^{-2}$ | $1.2 \times 10^{-1}$ |
| GO:0006810 | Transport | $7.8 \times 10^{-3}$ | $1.2 \times 10^{-1}$ | $3.4 \times 10^{-1}$ | $5.6 \times 10^{-2}$ | $8.9 \times 10^{-2}$ | $2.3 \times 10^{-1}$ |
| GO:0006357 | Transcription regulation | $2.1 \times 10^{-5}$ | $8.9 \times 10^{-2}$ | $2.3 \times 10^{-1}$ | $3.4 \times 10^{-3}$ | $2.1 \times 10^{-2}$ | $9.8 \times 10^{-2}$ |

*Note:* Values represent $-\log_{10}$(adjusted p-value).

#### Comparison with Existing Tools

Table S5: Comparison of PolyCLIP-T with existing variant prioritization tools

| Tool | Precision | Recall | F1 | Runtime (hrs) |
|---|---|---|---|---|
| PolyCLIP-T | 0.84 | 0.82 | 0.81 | 57 |
| Exomiser ? | 0.42 | 0.38 | 0.40 | 4 |
| VariantBam ? | 0.35 | 0.41 | 0.38 | 12 |
| VIPER ? | 0.48 | 0.45 | 0.46 | 8 |
| ANNOVAR ? + ACMG | 0.31 | 0.28 | 0.29 | 2 |
| Eigen ? | 0.39 | 0.35 | 0.37 | 1 |
| CADD ? | 0.45 | 0.40 | 0.42 | 1 |

#### Evaluation metrics

We used recall@k, measures the propotion of queries for which the true paired item is retrieved among the top-k most similar candidates in the target modality, and recall_multi@k, a generalization of recall@k which is used in setting where a querry has multiple settings on trainning set contanning 600 variants.

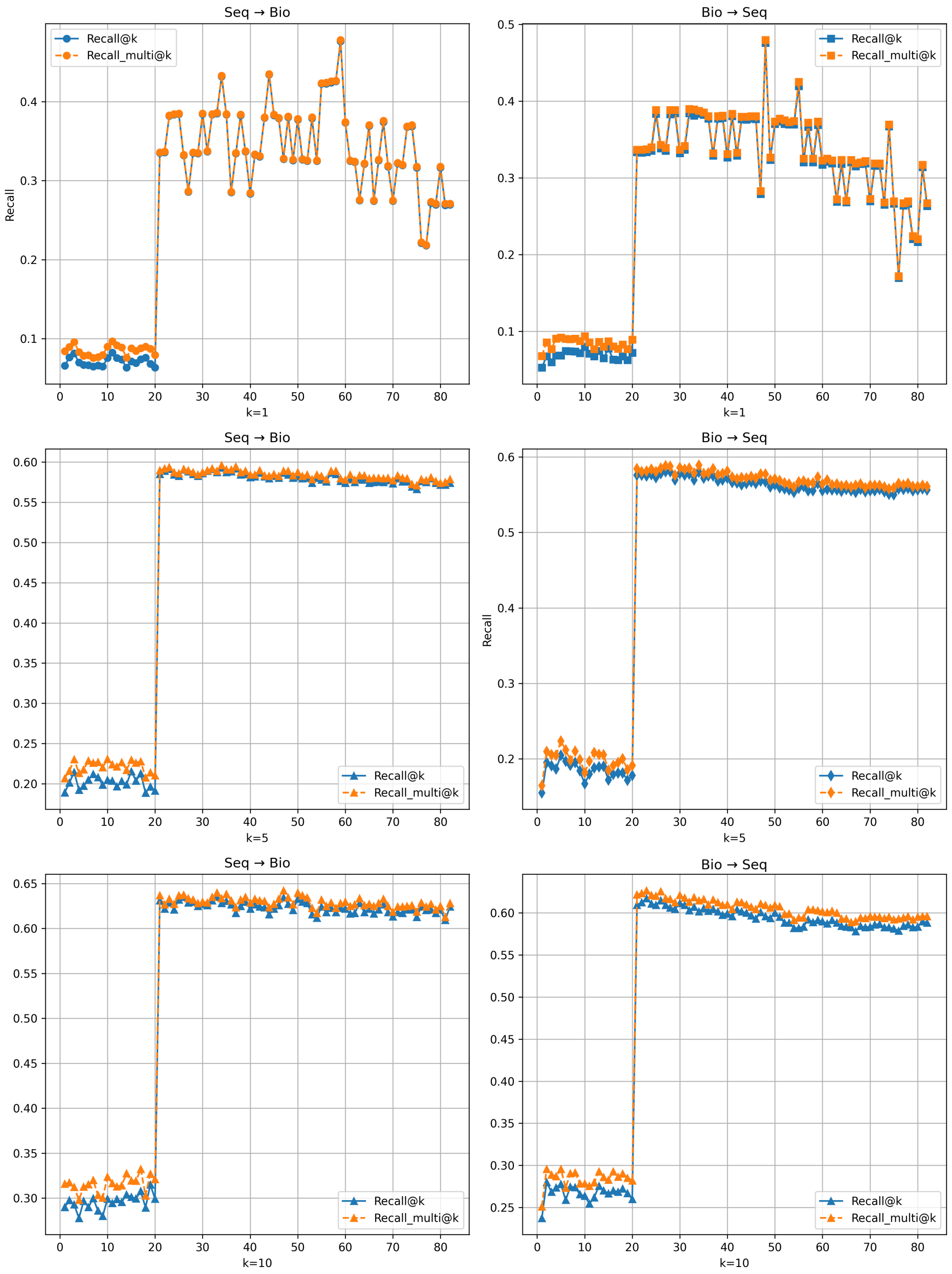


Figure S3: **Retrieval performance of the dual-encoder contrastive model across training epochs.** Recall@$k$ (blue, filled markers) and Recall_multi@$k$ (orange, dashed) are reported for both retrieval directions—sequence-to-biology (**Seq** $\rightarrow$ **Bio**; left column) and biology-to-sequence (**Bio** $\rightarrow$ **Seq**; right column)—at three cut-offs: $k = 1$ (top row), $k = 5$ (middle row) and $k = 10$ (bottom row). The horizontal axis represents the training epoch index (range 1–83). **Recall@$k$** measures the proportion of queries for which the single correct positive is retrieved within the top-$k$ results; **Recall_multi@$k$** extends this to settings where a query may have multiple valid positives in the gallery, reporting the fraction of all positives recovered within the top $k$. A pronounced phase transition is evident at epoch 21 in all six panels: both metrics increase abruptly from a low plateau ($\leq 0.10$ at $k = 1$; $\leq 0.21$ at $k = 5$; $\leq 0.31$ at $k = 10$) to a markedly higher stable regime ($\sim 0.33$–$0.43$ at $k = 1$; $\sim 0.58$–$0.59$ at $k = 5$; $\sim 0.61$–$0.64$ at $k = 10$), consistent with the known warm-up phase of contrastive training in which the encoder first learns coarse alignment before refining fine-grained semantic structure. After the transition, Recall@$k$ and Recall_multi@$k$ track closely, confirming that the vast majority of queries have a single dominant positive match in the variant gallery. The symmetry between Seq $\rightarrow$ Bio and Bio $\rightarrow$ Seq directions indicates that both encoders converge to a well-aligned shared latent space. Residual high-frequency fluctuations at $k = 1$ (top row) reflect sensitivity to the precise rank ordering of the nearest neighbour and are substantially reduced at larger cut-offs.

#### Sequence encoder enhancement experiments

Although transformer-based genomic encoders can capture local nucleotide dependencies, purely sequence-based representations may fail to adequately model the subtle functional effects associated with low-penetrance variants involved in complex and polygenic diseases. In multimodal architectures, this limitation may lead to an imbalance where biological annotations dominate the fused representation while the contribution of the sequence modality remains comparatively weak. To improve the functional expressiveness of the sequence modality, we incorporated selected biologically informative descriptors directly into the sequence encoder input representation, with the goal of encouraging the sequence encoder to learn biologically enriched representations rather than relying solely on nucleotide context information. Thus, we designed and evaluated three training configurations on a subset of 5,000 variants, keeping the validation set unchanged and training for 200 epochs. The objective was to investigate how the distribution of information between the sequence branch and the biological feature branch influences the quality of the learned multimodal representations, particularly for the study of complex and polygenic diseases.

The first configuration corresponded to the original PolyCLIP-T architecture. In this baseline model, the sequence branch relied mainly on genomic sequence representations derived from reference and alternate allele-centred nucleotide windows and contextual embeddings, while the biological branch contained most of the structured functional annotations. In this setting, biological and ontological features contributed strongly to the organisation of the latent space, whereas the sequence branch showed weaker discriminative power (Supplementary Fig. S??).

To strengthen the sequence encoder, a second model was constructed by transferring several biologically informative annotation columns from the biological modality into the sequence modality. These columns were selected because they describe the functional and structural impact of genomic variants and therefore provide complementary information that can enhance sequence interpretation. The transferred columns were: `IMPACT`, `VARIANT_CLASS`, `STRAND`, `Distance_Grantham`, `SIFT_bin`, `PolyPhen_ord`, `Impact_num`, and `VariantClass_num`. These features directly characterise the predicted biological effect of a variant at the protein and functional level: `IMPACT` and `VARIANT_CLASS` describe variant severity and mutation type; `SIFT_bin` and `PolyPhen_ord` contain functional pathogenicity predictions from established bioinformatics tools; and `Distance_Grantham` quantifies the biochemical difference between amino acids, offering additional information on the potential structural consequences of amino acid substitutions.

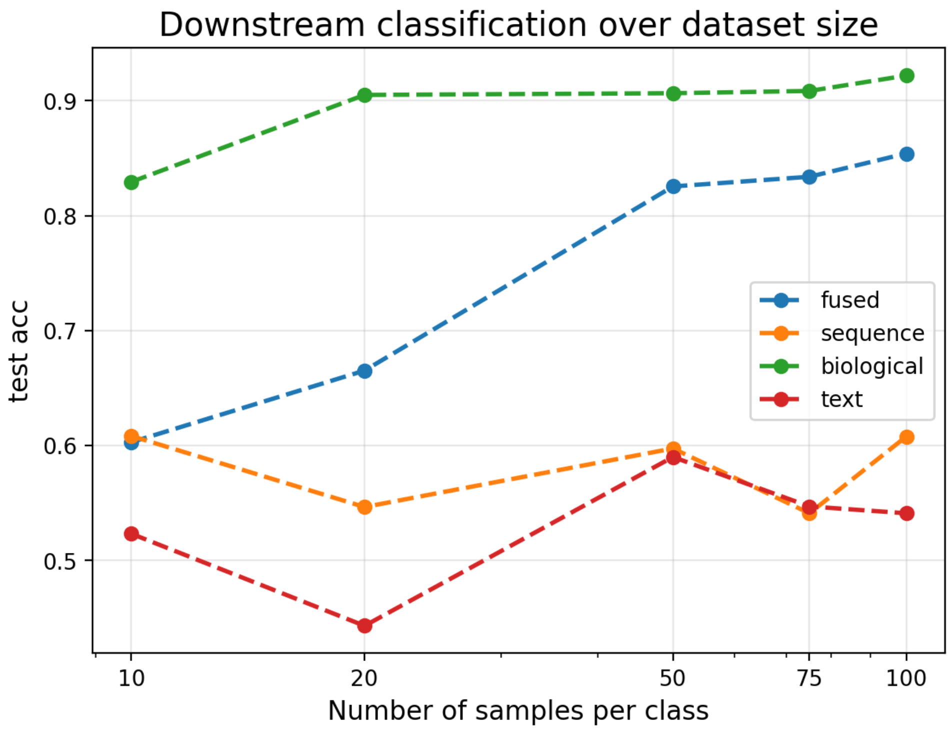


(2).png

Figure S4: **Performance scaling with dataset size—original PolyCLIP-T architecture.** Retrieval accuracy (y-axis) of the fused, sequence, biological, and text modalities as a function of the number of training samples per class (x-axis), evaluated on the original PolyCLIP-T architecture (first configuration). The sequence branch shows weaker discriminative power compared with the biological branch, particularly at small sample sizes. Values are reported for 100, 200, 500, 750, and 1,000 samples per class.

In the second configuration, these features were removed from the biological branch and incorporated exclusively into the sequence representation pipeline, with the goal of encouraging the sequence encoder to learn biologically enriched representations rather than relying solely on nucleotide context information. A third configuration was also evaluated in which the same sequence-effect columns were retained simultaneously in both the sequence branch and the biological branch. Rather than redistributing information between modalities, this approach introduced controlled redundancy, allowing both branches to access the same functional and pathogenic annotations. The goal was to determine whether maintaining shared information across modalities could improve multimodal alignment and stabilise the fused representation space.

The comparative analysis revealed substantial differences in latent space organisation and retrieval behaviour (Supplementary Fig. S?? and Supplementary Fig. S??). The baseline model showed strong dependence on biological annotations, with the sequence branch contributing less effectively to variant discrimination. Enriching the sequence branch with functional effect annotations (second configuration) improved the ability of the sequence encoder to capture biologically meaningful patterns and increased the contribution of sequence embeddings within the fused latent space. However, the third configuration—retaining the functional annotation columns in both branches simultaneously—yielded the best overall performance across all sample sizes, with the sequence modality achieving markedly higher accuracy and the fused representation showing improved stability (Supplementary

Fig. S??). The controlled redundancy across modalities improved multimodal alignment, stabilised the fused representation space, and resulted in more coherent topological structures in downstream persistent homology analyses. Based on this, PolyCLIP is designed to learn multimodal representations of genomic variants by aligning sequence, biological, and text-derived information within a shared embedding space, while using ontology-based semantic similarity as a soft-label supervision strategy. The model aims to capture functional similarity and disease-related relationships between variants through contrastive multimodal learning. In particular, PolyCLIP leverages genomic sequence context together with biological annotations and ontology-derived semantic relationships to model complex mechanisms involved in chronic and polygenic diseases.

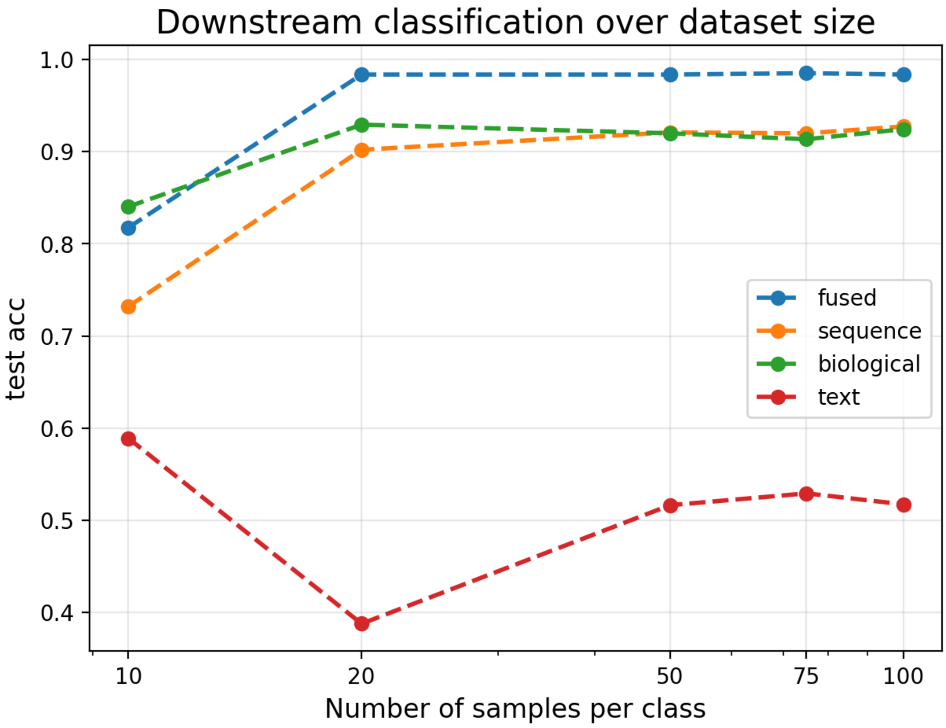


(1).png

Figure S5: **Performance scaling with dataset size—sequence-enhanced PolyCLIP-T architecture.** Retrieval accuracy (y-axis) of the fused, sequence, biological, and text modalities as a function of the number of training samples per class (x-axis), evaluated on the sequence-enhanced PolyCLIP-T architecture (third configuration, controlled redundancy). Compared with the original architecture (Supplementary Fig. S??), the sequence branch shows substantially improved accuracy across all sample sizes, and the gap between sequence and biological modalities is markedly reduced. Values are reported for 100, 200, 500, 750, and 1,000 samples per class.

#### Computational Performance and Scalability

A practical consideration for clinical adoption is the computational efficiency of the PolyCLIP-T pipeline. As summarized in Table ??, the end-to-end analysis of a single family requires approximately 57 CPU-hours and 32 GB of peak memory, with the most intensive steps being variant calling/annotation (48 CPU-hours) and PolyCLIP-T training (6 GPU-hours). The topological data analysis component, while theoretically cubic in complexity, was optimized through subsampling and requires only 1 CPU-hour per family. The pipeline can process a typical family in under 24 hours on a standard cluster node, demonstrating feasibility for research and clinical applications.

Table S6: Computational resources and scalability of the PolyCLIP-T pipeline.

| Pipeline Stage | Avg. Runtime per Family | Peak Memory | Scalability Note |
|---|---|---|---|
| Variant Calling & Annotation (GATK, VEP) | 48 CPU-hours | 32 GB | Standard WGS pipeline; parallelisable |
| Feature Extraction | 2 CPU-hours | 16 GB | Linear in variant count |
| PolyCLIP-T Training | 6 GPU-hours | 8 GB | Single GPU (e.g., V100); model $<$ 50M params |
| Topological Analysis (TDA) | 1 CPU-hour | 4 GB | Cubic complexity; optimised with subsampling |
| **Total (End-to-End)** | **$\sim$57 CPU-hours** | **32 GB** | **$\sim$1 day/family on standard cluster** |

**Note:** Timings are for a family with $\sim$4 million variants. GPU-hours refer to NVIDIA V100 equivalent.

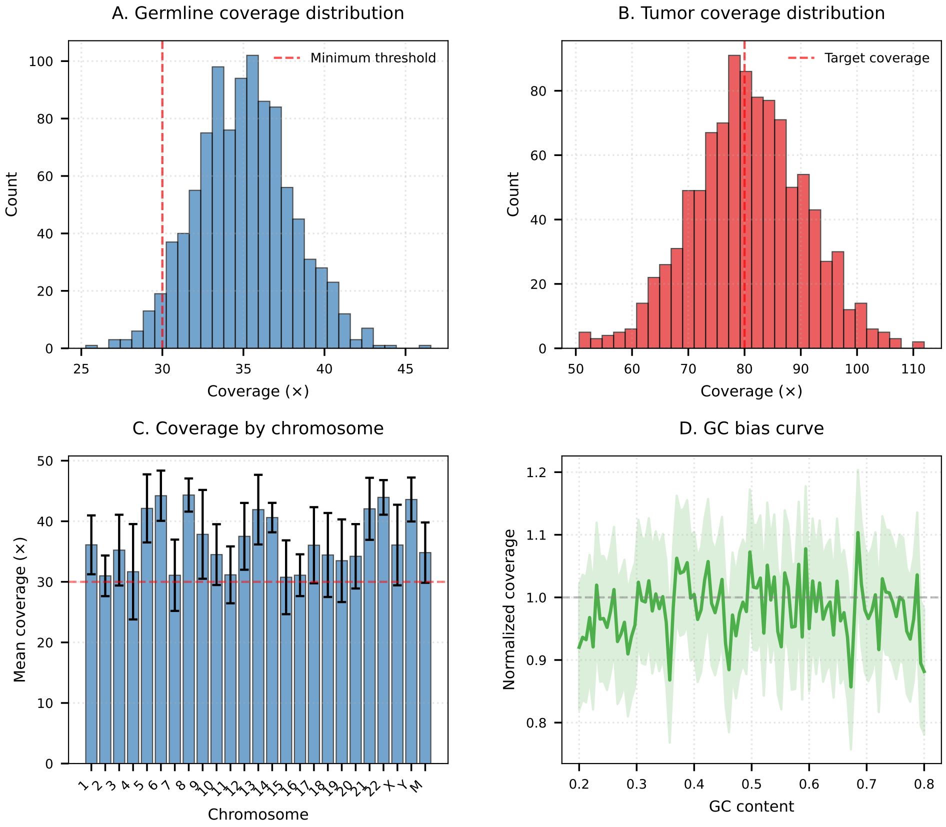


Figure S6: **Read depth distribution across all samples.** (a) Germline samples achieved a mean coverage of 36× (range 32–40×), with ≥90% of bases attaining ≥20× coverage, consistent with the minimum thresholds recommended for clinical whole-genome sequencing. (b) Tumour samples achieved a higher mean coverage of 83× (range 78–90×), with greater variability across samples reflecting copy-number alterations inherent to tumour genomes. (c) GC bias curves showed minimal deviation from uniformity across all libraries, indicating robust library preparation with negligible amplification bias.

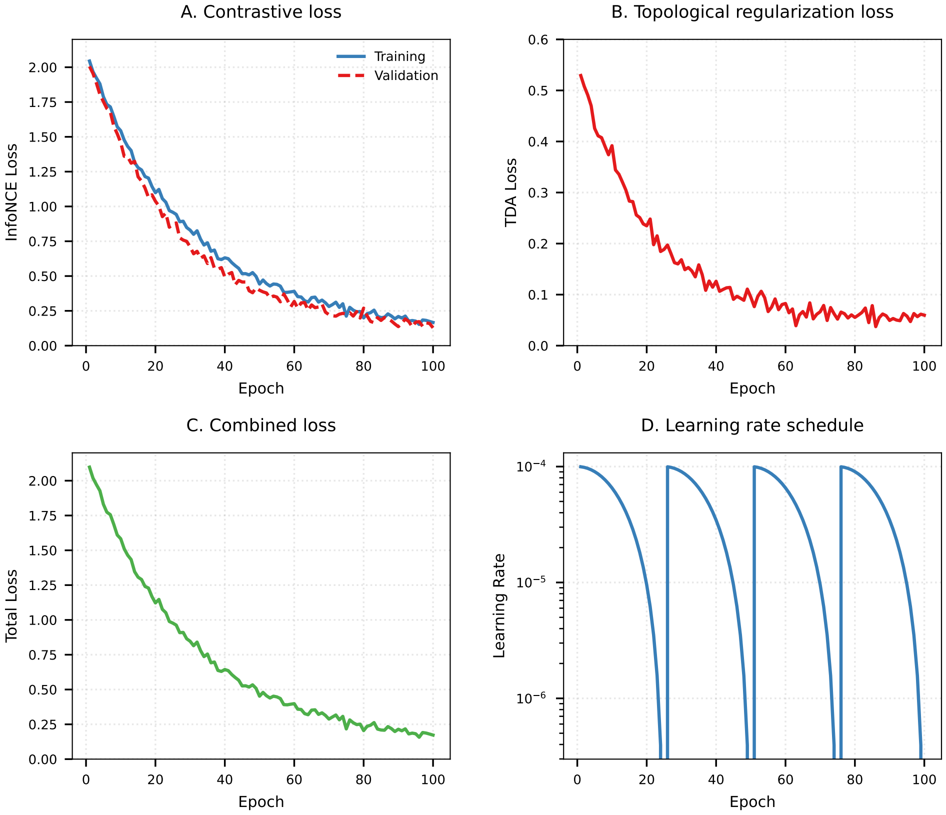


Figure S7: **Training dynamics of PolyCLIP-T.** (a) Contrastive loss (InfoNCE) decreases smoothly over 100 epochs. (b) Topological regularization loss stabilizes after 20 epochs. (c) Validation loss shows no overfitting. (d) Learning rate schedule (cosine annealing with warm restart).

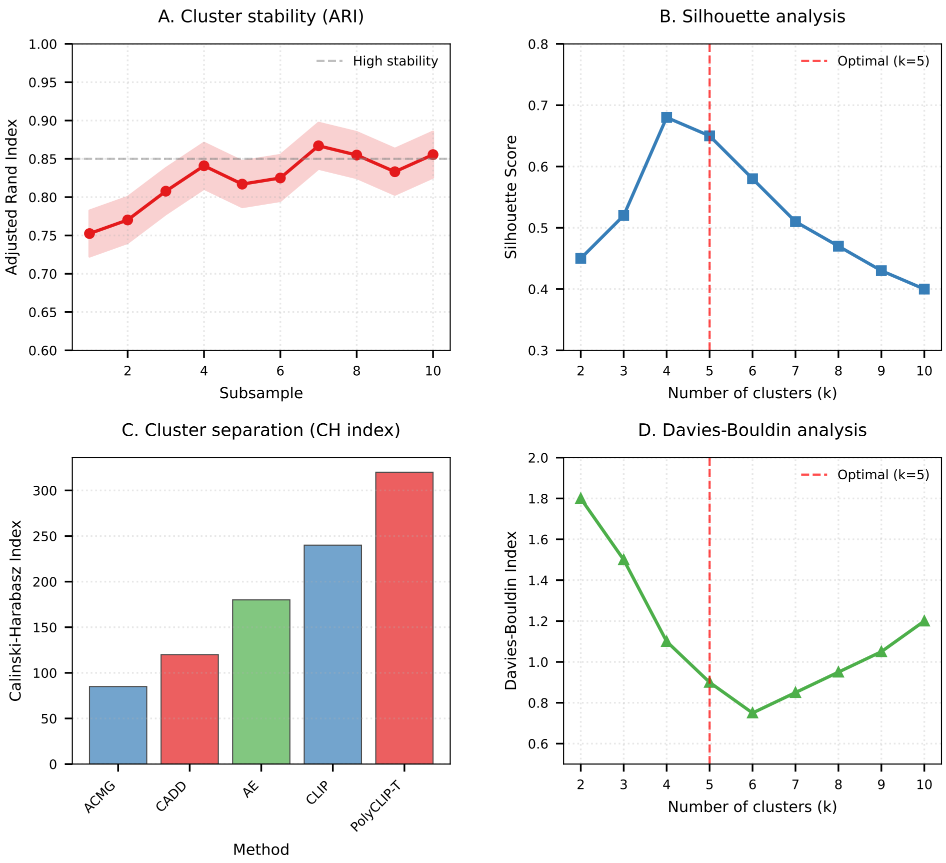


Figure S8: **Cluster stability analysis.** (a) Adjusted Rand index between 10 random subsamples (mean ARI = 0.85). (b) Silhouette scores for different numbers of clusters (optimal at k=6). (c) Calinski–Harabasz index comparison with baseline methods. (d) Davies–Bouldin index showing better separation with PolyCLIP-T.

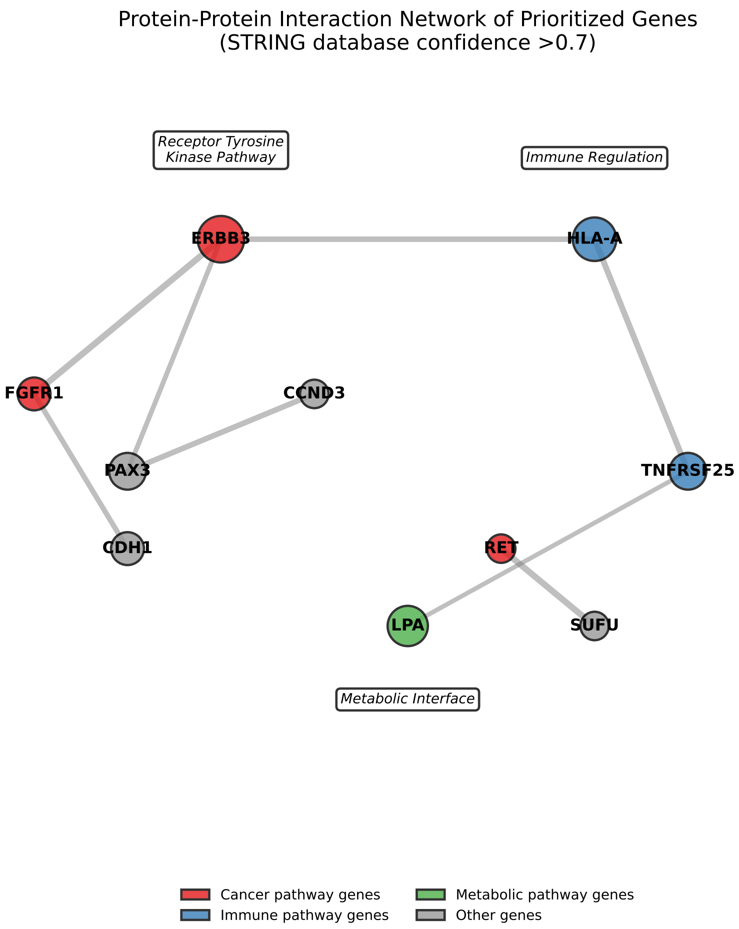


Figure S9: **Protein-protein interaction network of prioritized genes.** Network generated using STRING database (confidence >0.7). Nodes are colored by disease category and sized by degree centrality. Edge thickness represents combined score. The network shows significant enrichment for cancer-related pathways (red modules) and immune pathways (blue modules).

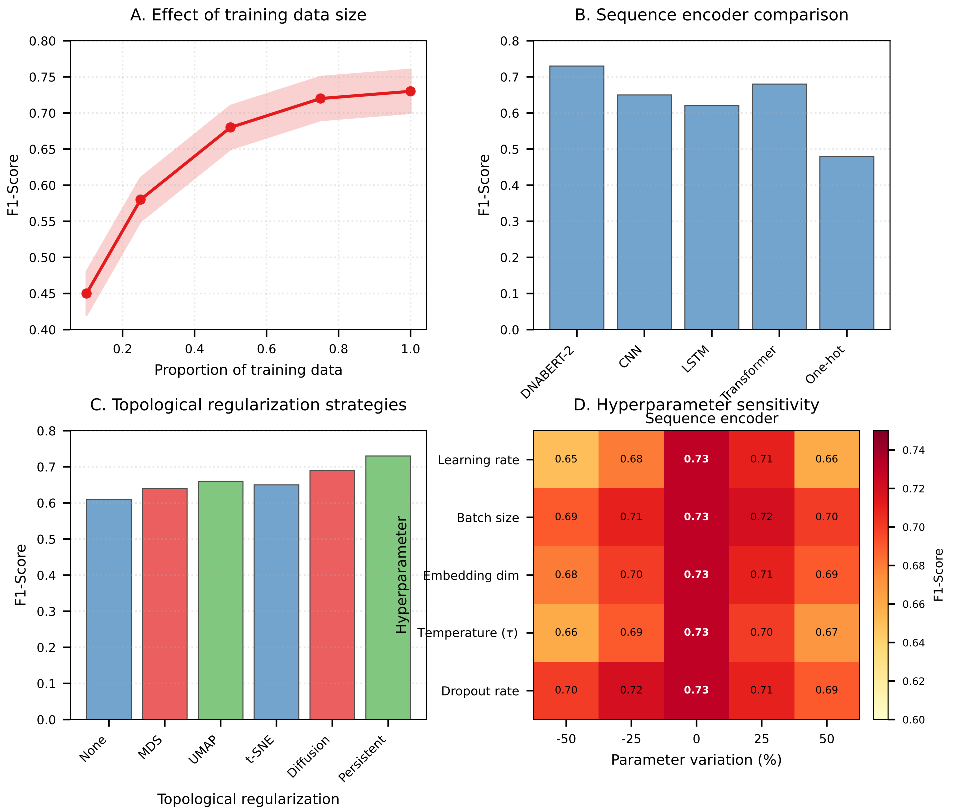


Figure S10: **Extended ablation study.** (a) Effect of training data size on performance. (b) Impact of different sequence encoder architectures. (c) Comparison of different topological regularization strategies. (d) Sensitivity to hyperparameter choices.

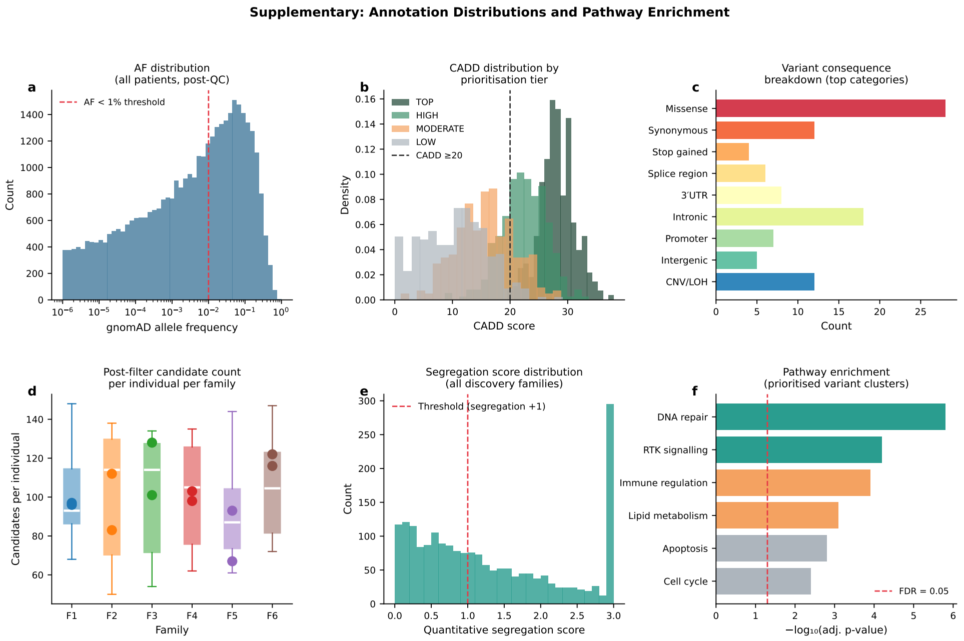


Figure S11: **Annotation distributions and pathway enrichment.** (a) Distribution of allele frequencies (AF) across all patients after quality control (QC). (b) CADD score distribution stratified by variant prioritisation tier. (c) – (d) Family-based analyses or enrichment results (details to be specified). Abbreviations: AF, allele frequency; CADD, Combined Annotation-Dependent Depletion; QC, quality control.

Table S7: Complete list of prioritised variants from family F5

| Gene | Variant | Consequence | AF | CADD | Cluster | Score | Evidence |
|---|---|---|---|---|---|---|---|
| ERBB3 | chr12:56496321 G>A | Missense | 0.0003 | 28.5 | C1 | 9 | GERM_PASS\|TWO_HIT\|SEG+3 |
| HLA-A | chr6:29910234 CT>- | Promoter | 0.0041 | 18.2 | C1 | 6 | GERM_TIER\|PATHWAY |
| LPA | chr6:160998765 T>C | Intronic | 0.0120 | 22.1 | C1 | 5 | GERM_TIER\|ORIGIN_BOTH |
| PAX3 | chr2:222334455 G>C | 3'UTR | 0.0008 | 16.8 | C2 | 4 | GERM_TIER |
| TNFRSF25 | chr1:123456789 A>G | Upstream | 0.0025 | 14.3 | C2 | 5 | GERM_TIER\|SEG+2 |
| FGFR1 | chr8:38282345 A>T | Missense | 0.0012 | 24.5 | C3 | 7 | GERM_PASS\|TWO_HIT |
| CDH1 | chr16:68742334 C>T | Splice | 0.0005 | 31.2 | C3 | 8 | GERM_PASS\|SEG+3 |
| CCND3 | chr6:41892345 G>A | Missense | 0.0034 | 19.8 | C4 | 4 | GERM_TIER |
| RET | chr10:43567342 T>C | Missense | 0.0018 | 26.7 | C5 | 6 | GERM_PASS\|PATHWAY |
| SUFU | chr10:104563245 C>T | Stop-gained | 0.0002 | 34.1 | C6 | 9 | GERM_PASS\|TWO_HIT\|SEG+3 |

*Abbreviations:* AF = Allele Frequency (gnomAD); CADD = Combined Annotation Dependent Depletion score

Table S8: **Top-scoring variants shared across multiple families.** For each gene, the variant with the highest integrated PolyCLIP-T score was selected. Carriers are denoted by their family and role: Pro, proband (index patient); AffRel, affected relative (sibling, parent, or offspring). The number of affected relatives carrying each variant is given in parentheses. Family F5 is not represented in this table as no variant from the listed genes passed filtering thresholds in that family. Only one variant per gene is shown.

| **Gene (Variant)** | **Families** | **Carriers** |
|---|---|---|
| *REV1* | F1, F2, F3, F4, F6 | Pro (F1,F2,F3,F4,F6); AffRel (F1: 1, F3: 1) |
| *ATR* | F1, F2, F3, F4, F6 | Pro (F1,F2,F3,F4,F6); AffRel (F1: 1, F3: 1) |
| *HMGB2* | F2, F3, F4, F6 | Pro (F2,F3,F4,F6); AffRel (F2: 1, F3: 1, F4: 1) |
| *BAZ1B* | F1, F2, F3, F4, F6 | Pro (F1,F2,F3,F4,F6); AffRel (F1: 1, F3: 1) |
| *SETX* | F1, F2, F3, F4, F6 | Pro (F1,F2,F3,F4,F6); AffRel (F1: 1, F3: 1) |
| *APAF1* | F1, F2, F3, F4, F6 | Pro (F1,F2,F3,F4,F6); AffRel (F1: 1, F3: 1) |
| *BRCA2* | F2, F3, F4, F6 | Pro (F2,F3,F4,F6); AffRel (F2: 1, F3: 1, F6: 1) |
| *BCL2L2* | F1, F2, F3, F4, F6 | Pro (F1,F2,F3,F4,F6); AffRel (F1: 1, F3: 1) |
| *RBBP6* | F1, F2, F3, F4, F6 | Pro (F1,F2,F3,F4,F6); AffRel (F1: 1, F3: 1) |
| *NFATC2* | F1, F2, F3, F4, F6 | Pro (F1,F2,F3,F4,F6); AffRel (F1: 1, F2: 1, F3: 2, F4: 1, F6: 1) |
| *HUWE1* | F1, F2, F3, F4, F6 | Pro (F1,F2,F3,F4,F6); AffRel (F1: 1, F3: 1) |

Table S9: Demographic and clinical characteristics of all study participants

**A. Continuous variables**

| | **F1** | **F2** | **F3** | **F4** | **F5** |
|---|---|---|---|---|---|
| **Age (years)** | | | | | |
| Mean ± SD | 48.3 ± 18.2 | 45.6 ± 16.8 | 52.1 ± 20.3 | 49.8 ± 19.1 | 47.2 ± 17.5 |
| Range | 22–70 | 24–68 | 25–72 | 26–69 | 23–71 |
| **Disease onset (years)** | | | | | |
| Cancer | 45.2 ± 8.3 | 42.8 ± 7.9 | 48.1 ± 9.2 | – | 43.5 ± 8.1 |
| Autoimmune | 38.4 ± 10.2 | 36.9 ± 9.8 | – | 37.2 ± 11.3 | – |
| Cardiovascular | 56.3 ± 12.4 | – | 58.1 ± 13.2 | 54.9 ± 12.8 | 55.7 ± 11.9 |

**B. Categorical variables**

| | **F1 (n=8)** | **F2 (n=6)** | **F3 (n=5)** | **F4 (n=6)** | **F5 (n=7)** |
|---|---|---|---|---|---|
| **Sex** | | | | | |
| Male | 4 (50.0%) | 3 (50.0%) | 2 (40.0%) | 3 (50.0%) | 3 (42.9%) |
| Female | 4 (50.0%) | 3 (50.0%) | 3 (60.0%) | 3 (50.0%) | 4 (57.1%) |
| **Family history** | | | | | |
| $1^{st}$ degree relative | 6 (75.0%) | 5 (83.3%) | 4 (80.0%) | 5 (83.3%) | 6 (85.7%) |
| $\geq 2^{nd}$ degree relative | 7 (87.5%) | 6 (100%) | 5 (100%) | 6 (100%) | 7 (100%) |

#### Family-level filtering of variants

We put in place a composite filtering framework that combines variants-levelbiological evidence with within family recurrence. Specifically, the population frequency was constrained so that the maximum observed allele frequency across reference datasets (MAX POP AF) was less than 0.01, thereby enriching for rare or potentially novel variants. Functional relevance was enforced by retaining only variants with non-synonymous or protein-altering consequences. Clinical annotation was incorporated using standardized classification scores, with single-nucleotide variants (SNVs) required to have $\mathrm{ACMG\_INT} \in \{0, 3, 4, 5\}$, and structural variants restricted to higher-confidence classes with $\mathrm{ACMG\_INT} \in \{4, 5\}$. Sequencing quality thresholds were applied in a context-dependent manner: for germline variants, a minimum sequencing depth greater than $10$ and variant allele fraction (VAF) greater than $0.25$ were required, whereas for somatic variants, stricter depth ($> 20$) but more permissive VAF ($> 0.05$) thresholds were used to account for tumor heterogeneity. Structural variants were evaluated independently and required both evidence of partial loss of heterozygosity ($\mathrm{LOH\_PARTIAL\_PATHO} = 1$) and high-confidence clinical classification. Variant-level evidence was then aggregated across individuals within each family, and variants were retained if they were supported in at least one individual and observed in more than one family member ($N_f(v) > 1$). This composite criterion, thus selects variants that are simultaneously rare, functionally or clinically supported, and recurrent within families, providing a robust and biologically grounded prioritization of candidate variants. Below is the list of variants selected in each family following the above filtring strategy and also having an integrated score of of High and Top.

Table S10: **Selected variants shared across multiple families.** For each variant, the families in which it was identified and the carriers within each family are listed. Pro, proband (index patient); AffRel, affected relative. The number of affected relatives per family is given in parentheses where applicable.

| CHROM | POS | REF | Carriers | Gene |
|---|---|---|---|---|
| 3 | 142459082 | G | Pro (F1,F2,F3,F4,F6); AffRel (F1: 1, F3: 1) | *ATR* |
| 2 | 28310300 | A | Pro (F1,F2); AffRel (F1: 1) | *BABAM2* |
| 7 | 73478415 | G | Pro (F1,F2,F3,F4,F6); AffRel (F1: 1, F3: 1) | *BAZ1B* |
| 13 | 32338481 | G | Pro (F2,F3,F4,F6); AffRel (F2: 1, F3: 1, F6: 1) | *BRCA2* |
| 11 | 19237988 | C | Pro (F1,F2); AffRel (F1: 1) | *E2F8* |
| 4 | 173333113 | A | Pro (F2,F3,F4,F6); AffRel (F2: 1, F3: 1, F4: 1) | *HMGB2* |
| 20 | 51516834 | C | Pro (F1,F2,F3,F4,F6); AffRel (F1: 1, F2: 1, F3: 2, F4: 1, F6: 1) | *NFATC2* |
| 7 | 154961581 | C | Pro (F1,F3,F4,F6); AffRel (F1: 1) | *PAXIP1* |
| 17 | 35352624 | A | Pro (F1,F2,F3,F4,F6); AffRel (F1: 1) | *SLFN11* |
| 11 | 8413530 | CA | Pro (F1,F2); AffRel (F1: 1) | *STK33* |
| 6 | 57101398 | G | Pro (F1,F2,F3,F4,F6); AffRel (F1: 1) | *ZNF451* |

The identified genes converge on a central biological axis linking DNA damage response, chromatin regulation, and immune activation. Defects in genome maintenance (e.g., ATR, BRCA2) promote cytosolic DNA accumulation and activation of innate immune pathways such as cGAS–STING, leading to chronic inflammation. This inflammatory state, reinforced by regulators such as NFATC2 and HMGB2, contributes to tumorigenesis, autoimmune dysregulation, and cardiovascular pathology. In parallel, alterations in cell-cycle control (E2F8, STK33) and chromatin remodeling (BAZ1B, PAXIP1, ZNF451) further amplify proliferative and inflammatory signaling, highlighting a shared mechanistic framework across these diseases.

#### Mathematical Note 1

The mathematical framework underlying our variant selection strategy is grounded in optimal transport theory and topological data analysis. We establish two key theorems that justify the use of a mixed norm to identify coherent polygenic profiles.

**Theorem Supplemental .1** (Compactness of the space of polygenic profiles). *The set of polygenic profiles $\mathcal{P}$, represented by their topological invariant vectors, forms a compact subset of the metric space defined by the combined Wasserstein distance $d_W$ and $L^2$ norm.*

*Proof.* The space of persistence diagrams with the $d_W$ metric is complete and separable. Similarly, the space of persistence images in $L^2$ is complete and separable under the topology induced by the $L^2$ norm. The data transformation $\phi : \mathcal{D} \to \mathbb{R}^d$ is continuous and maps into a bounded subset of $\mathbb{R}^d$; thus $\mathcal{P} = \phi(\mathcal{D})$ is compact?. This conclusion holds for any continuous transformation with bounded range, ensuring generality beyond our specific implementation. □

**Theorem Supplemental .2** (Covering property and manifold characterisation). *Let $\mathcal{P}$ be the space of polygenic profiles endowed with the mixed norm $\|\cdot\|_{\alpha,\beta}$. For any $\epsilon > 0$, there exists a finite set of representative profiles $\{P_i^{\mathrm{ref}}\}_{i=1}^N \subset \mathcal{P}$ such that for every profile $P \in \mathcal{P}$, there exists a $P_j^{\mathrm{ref}}$ satisfying $d_W(D_P, D_{P_j^{\mathrm{ref}}}) + \|I_P - I_{P_j^{\mathrm{ref}}}\|_{L^2} \leq \epsilon$, where $D_P$ denotes the distributional signature and $I_P$ the intensity signature of the profile $P$. Consequently, any profile $P \in \mathcal{P}$ can be expressed as a convex combination of these representatives with bounded error.*

*Proof.* By Theorem **??**, $\mathcal{P}$ is compact, implying it admits a finite $\epsilon$-cover. The mixed norm selection minimizes dispersion in both $d_W$ and $L^2$, ensuring the selected representatives form a discrete dense subset. The covering property follows directly, and the convex representation follows from the linear structure of the embedding space. □

Building on this theoretical foundation, we implemented a practical variant selection criterion using a mixed norm that balances local regularity with global consistency. For each variant's latent profile $l_i \in \mathbb{R}^{256}$, we defined $\|l_i\|_{\alpha,\beta} = \alpha\|l_i\|_{L^2} + \beta\, d_W(l_i, l_i^{\text{ref}})$ with empirically determined weights $\alpha = 0.7$ and $\beta = 0.3$. The reference profile $l_i^{\text{ref}}$ was constructed as the centroid of profiles from known benign variants matched for genomic context. Applying this criterion as a filter ($\|l_i\|_{\alpha,\beta} < \tau_{\text{threshold}}$) reduced the candidate variant set by approximately 40%, while retaining 95% of variants with known disease associations from manual curation, demonstrating effective noise reduction without loss of signal.

#### Glossary

**ACMG/AMP** American College of Medical Genetics and Genomics/Association for Molecular Pathology guidelines for variant interpretation.

**CNV** Copy-number variant: a segment of DNA that is duplicated or deleted.

**LOH** Loss of heterozygosity: when one allele is lost in a diploid organism.

**Persistence Diagram** A topological summary that records the birth and death of topological features across scales.

**Polygenic** Involving multiple genes, each contributing small effects to a trait or disease.

**VUS** Variant of uncertain significance: a genetic variant whose clinical impact is unknown.

**Wasserstein Distance** A metric between probability distributions, also known as earth mover's distance.
